# Restoring STAR*D: A RIAT Reanalysis of Medication Augmentation Therapy After Failed SSRI Treatment Using Patient-Level Data with Fidelity to the Original Research Protocol

**DOI:** 10.1101/2025.10.27.25338365

**Authors:** Colin Xu, Thomas T. Kim, Martin Plöderl, Kevin P. Kennedy, Irving Kirsch, Jay D. Amsterdam, H. Edmund Pigott

## Abstract

**Background:** The STAR*D trial is the most influential study of sequential antidepressant treatment strategies. However, major STAR*D publications deviated from the protocol-defined analytic plan. Prior re-analyses found lower cumulative remission rates than STAR*D publications reported, sustained remission rates of only 3.1 to 8.4% at 12 months, and high rates of treatment-emergent suicidal ideation (TESI) during medication-switch therapy. A similar reanalysis is warranted for STAR*D’s augmentation study in which citalopram was augmented with sustained-release bupropion or buspirone.

**Methods:** We reanalyzed STAR*D’s patient-level augmentation dataset with fidelity to the original protocol or relevant STAR*D publications where the protocol did not prespecify an analytic plan.

**Results:** This reanalysis identified 124 patients (21.9% of enrolled subjects) who were inappropriately included in the original STAR*D analysis, including 54 who were in protocol-defined remission before starting augmentation therapy. Remission rates as defined in the protocol were lower than reported in the original publication for bupropion SR (25.0% vs 29.7%) and buspirone (25.8% vs. 30.1%). Using a secondary definition of remission, bupropion SR’s rate was significantly lower than reported in original publications (29.2% vs. 39.0%). Sustained remission through 12 months was low (4.9–12.5%). TESI rates were significantly higher for buspirone (13.9%) than bupropion SR (3.6%) augmentation.

**Conclusion:** Compared with the original STAR*D publication, our reanalysis identified inflated remission rates, low sustained remission, and marked differences in TESI risk between augmentation strategies. These findings suggest that both treatments offer lower acute and sustained benefit than is widely understood, with buspirone associated with more TESI.

## Introduction

The NIMH-funded *Sequenced Treatment Alternatives to Relieve Depression* (STAR*D) trial is the largest and most influential treatment study ever conducted of outpatients with major depressive disorder (MDD), with over 100 journal articles published by study investigators.^1-7^ However, previous research by our group raised concerns about deviations from the protocol in original publications, primarily the use of a post-hoc remission outcome measure and inappropriate inclusion of subjects in the outcomes analysis. Our previous Restoring Invisible and Abandoned Trials (RIAT) re-analyses of STAR*D found markedly lower remission rates, low long-term sustained remission rates, and high rates of treatment-emergent suicidal ideation (TESI).^8,9^

Briefly, STAR*D provided up to four consecutive treatment trials per patient and was designed to give guidance in selecting the best next-step treatment for the patients who fail to remit from their initial and/or subsequent treatments. To comport with real-world clinical practice, the STAR*D investigators employed an open-label pragmatic trial design that only enrolled patients seeking routine medical or psychiatric care and included patients with a wide range of comorbid medical and psychiatric conditions.

A total of 4041 patients screened positive for MDD and were enrolled. Of these patients, 3110 met STAR*D’s inclusion for data analysis criteria which was a score >13 on the blindly-administered 17-item Hamilton Rating Scale for Depression (HRSD).^1,Figure 1^ In step-1, all patients received citalopram monotherapy. Patients who failed to remit with (or were intolerant of) citalopram could select other treatment options for randomization in step-2.

**Figure 1.**
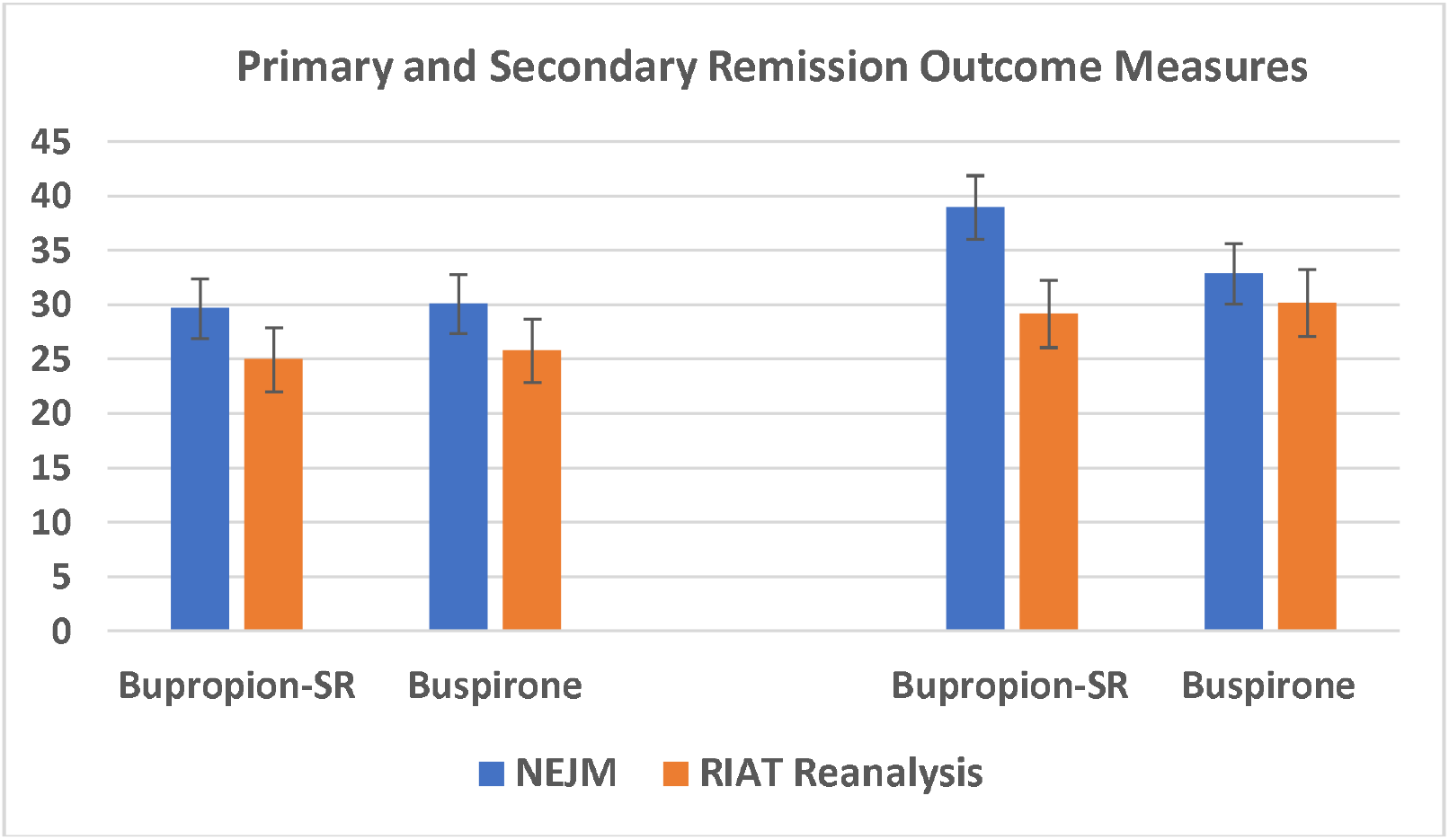
Comparison of the Primary and Secondary Outcome Measures for Remission. Error bars represent standard errors of the proportion.

In step-2, patients had seven treatment options to choose from: four switch options in which citalopram was stopped and the new treatment started (one of three antidepressants or cognitive therapy [CT]) and three augmentation options in which citalopram was combined with an additional treatment (one of two medications or CT). The medications used for augmentation in step-2 were sustained-release bupropion (bupropion SR) and buspirone, “*two commonly used augmentation agents with distinct pharmacologic profiles*.” ^3,p.1244^.

Despite being encouraged to accept randomization to all seven treatment options, only 1.5% of patients did so, with the vast majority opting to either have their medication switched or augmented with a second medication.^2,3,10^ Too few patients accepted the possibility of being assigned to CT so it was excluded from STAR*D’s primary analyses.^2,3^

The STAR*D investigators reported that patients who opted to switch their medication were significantly more depressed entering step-2 and had higher rates of side effects with citalopram in step-1 than patients who opted to augment citalopram with another medication.^2^ Given these differences, the investigators analyzed the medication-switch and medication-augmentation groups separately in the published articles.^2,3^

The STAR*D investigators reported no significant differences in remission or response rates between the two augmentation treatments. However, they did note that bupropion had a significantly lower dropout rate due to medication intolerance compared to buspirone (12.5% vs. 20.6%). Bupropion was also associated with a greater reduction in scores on the Quick Inventory of Depressive Symptomatology– Self-Report (QIDS-SR), a patient self-report scale administered during clinic visits.^3^

However, using the QIDS-SR as outcome was an undeclared deviation from the NIMH-protocol, which explicitly prohibited the use of clinic-administered assessments like the QIDS-SR and its clinician-administered version (QIDS-C) for reporting research outcomes. This was because these assessments were not blinded and only used to guide care.^11,12^ The results as planned in the protocol were never published by the STAR*D investigators.

A reanalysis of STAR*D’s step-2 augmentation dataset is warranted for three reasons:

First, our two prior reanalysis efforts under the RIAT initiative discovered protocol violations which led to an overestimation of remission rates.^8,9^ Similar discrepancies could be present in STAR*D’s step-2 augmentation treatments and therefore should be explored.

Second, the 12-month follow-up results of the augmentation therapies remain unpublished despite STAR*D’s primary investigator stating in 2007 that STAR*D investigators would “*compare longer-term outcomes of the various treatments*.”^13, p.201^

Third, the occurrence of TESI during step-2 augmentation treatments remains unpublished. Our reanalysis of STAR*D’s medication-switch therapies found a significant increase in TESI during step-2 treatment compared to step-1 citalopram therapy that averaged 43% (12.9% vs 9.0%).^9^ This is important, as an analysis of the Food and Drug Administration Adverse Event Reporting System found that medications associated with TESI are also linked to increased suicide attempts and suicides.^14^

Meaningful differences in the likelihood of patients experiencing TESI during augmentation treatment, as well as the sustainability of treatment gains, are critical information in the decision-making process for physicians and patients and therefore vital for them in determining the best next-step treatment option after a failed response to initial SSRI therapy.

The current RIAT study reanalyzed STAR*D’s step-2 augmentation trial with fidelity to the original STAR*D research protocol and related publications. In addition to using STAR*D’s primary outcome (i.e., the Hamilton Rating Scale for Depression [HRSD]), we also compared: (a) outcomes for patients who entered the one-year follow-up phase; and (b) TESI rates using the same methodology STAR*D investigators used in a secondary analysis to identify patients who became suicidal during citalopram treatment.^15^

## Methods

### Data

We used STAR*D’s patient-level dataset downloaded from NIMH in January 2025. We reference the 2006 STAR*D publication in *The New England Journal of Medicine* (NEJM) when discussing the published STAR*D data.^3^

### Subjects

Patients enrolled in the STAR*D trial were 18 to 75 years old, diagnosed with nonpsychotic MDD, and seeking care at 18 primary and 23 psychiatric care clinical sites across the United States.

We adhered to STAR*D’s exclusion from data analysis criteria. We excluded patients with a baseline HRSD score <14 or missing a HRSD score at entry into step-1 ^(see:1, figure 1)^ as well as those who had an HRSD score <8 at entry into step-2 since the STAR*D investigators prespecified that “*patients who begin a level with HRSD <8 will be excluded from analyses*” ^16, p.130^

Supplement 1 is the patient flow chart moving from step-1 citalopram treatment to the step-2 medication-augmentation therapies.

### Original STAR*D treatment and assessment of outcomes

STAR*D investigators sought to provide the highest quality of acute and continuing-care treatment to maximize the number of remissions while minimizing the likelihood of relapse and dropouts (see Supplement 2). Each of the four treatment steps consisted of 12 weeks of antidepressant therapy, with an additional 2 weeks for patients deemed close to remission. All patients received citalopram as their step-1 treatment.

All treatments were administered using a system of measurement-based care that assessed depressive symptom severity using the QIDS-SR and QIDS-C at baseline and at weeks 2, 4, 6, 9, and 12 to guide medication dosing. Patients without a satisfactory response (or significant intolerance) to citalopram in step-1 had the option to enter step-2 and either switch or augment their citalopram therapy.

The STAR*D investigators implemented an “equipoise-stratified randomized” research design, whereby patients could accept randomization to only certain step-2 treatment strategies, such as medication-augmentation or medication-switch.^17^ Among their accepted strategies, patients were then randomized to a specific treatment. While patients were strongly encouraged to accept randomization into any of the seven step-2 treatments options, only 4.0% of patients accepted randomization to any medication-switch and any medication-augmentation options, thereby preventing a head-to-head comparison between medication-switch and medication-augmentation treatments.^2^

In the step-2 augmentation therapies, patients who were randomized to augmentation continued their established citalopram dose and added either bupropion augmentation (starting at a daily maximum dose of 200 mg, with the potential to increase to 400 mg total daily dose); or buspirone augmentation (starting at a daily maximum dose of 15 mg, with the potential to increase to 60 mg total daily dose) (see Supplement 3 for a description of the medications and their dosing).

### Primary STAR*D outcome of symptom remission

For the current RIAT reanalysis we used STAR*D’s protocol-stipulated primary outcome, which was remission defined as a posttreatment HRSD score <8. The HRSD was obtained at treatment entry and exit by research outcome assessors (ROAs) blind to treatment status. In their publications, STAR*D investigators prespecified in step-1 that “*primary analyses classified patients with missing exit HRSD scores as nonremitters a priori*”^1, p.34^ and similarly stated in step-2, “*patients who did not undergo HRSD-17 evaluation at the end of the study were declared a priori not to have had a remission*.”^2, p.1238^ In our RIAT reanalysis, we analyze the primary outcome data using these prespecified criteria.

Due to the high rate of augmentation patients missing an exit HRSD (n = 110; see Supplement 4), we conducted a sensitivity analysis in which we imputed HRSD scores by converting their final QIDS-SR score to an HRSD score. We then estimated secondary outcome measures for remission, response (defined as a ≥50% reduction in HRSD), and mean symptom improvement on the HRSD. This is the same imputation method used by the STAR*D investigators.^2^

### TESI Analysis

In a secondary analysis, STAR*D investigators found that some patients in step-1 experienced TESI after citalopram initiation.^15^ TESI was defined as a score of ≥2 on any postbaseline visit for patients whose baseline TESI score was <2 on the QIDS-C suicide item, which was scored as the following: “(0) *Does not think of suicide or death;* (1) *Feels life is empty or is not worth living;* (2) *Thinks of suicide/death several times a week for several minutes; and;* (3) *Thinks of suicide/death several times a day in depth, or has made specific plans, or attempted suicide*.”^18^ For our RIAT reanalysis, we use these same criteria to analyze TESI with the baseline score being at entry into step-2.

### One-year follow-up treatment for responders and remitters

According to the study plan, patients scoring <6 on their last clinic-administered QIDS-C during step-2 treatment were encouraged to enter the 12-month follow-up phase. A QIDS score <6 was considered by STAR*D investigators to correspond to a HRSD score <8, STAR*D’s definition of remission.^19^ Clinicians strongly encouraged patients who did not obtain a QIDS-C-defined remission to enter the next-step treatment. Additionally, patients who responded but did not remit on the QIDS-C and who declined randomization to a next-step treatment were encouraged to enter the 12-month follow-up, too.

During follow-up, patients continued their “*previously effective acute treatment medication(s) at the doses used in acute treatment but that any psychotherapy, medication, or medication dose change could be used to sustain benefit*.”^7, p.1908^

During the follow-up phase, the STAR*D ROAs collected HRSD data from patients at months 3, 6, 9, and A telephonic integrated voice response version of the QIDS-SR (QIDS-IVR) was also collected at months 1, 2, 4, 5, 7, 8, 10, and 11. In order to gain the most complete follow-up dataset, we imputed missing quarterly HRSD scores by mapping the closest available QIDS-IVR score ± 4 weeks to that visit timepoint.

In that STAR*D’s protocol was silent on how to analyze the follow-up data, we defined relapse as an HRSD score ≥14 at any one of the four follow-up timepoints (months 3, 6, 9, and 12). For patients who met remission criteria and entered follow-up, we calculated the number of patients who sustained their remission (i.e., HRSD score <8 at all four timepoints). Similar to the STAR*D investigators in their Texas Medication Algorithm Project (T-MAP) article, we also used a “lenient” definition of sustained remission irrespective of missing data at some timepoints.^20^

### Statistical analyses

Augmentation treatments were compared with χ^2^-tests for discrete outcomes (remission, response, TESI, relapse and sustained remission) and ANOVA for continuous outcomes. Results of our RIAT reanalysis of step-2 remission rates were compared against those reported in the original STAR*D publication using χ^2^ tests.^3^

We used R version 4.4 for all analyses.

Supplement 5 provides the statistical code that the first and second authors used to analyze the STAR*D patient-level data from the NIMH-provided datasets.

## Results

### Patient characteristics

In the current RIAT reanalysis, 441 augmentation patients met the inclusion criteria for data analysis. Table 1 presents the number of augmentation patients who were excluded in our reanalysis (n = 124), yet included in the STAR*D investigators’ step-2 article, and the reasons for their exclusion.

**Table 1:** Number of Augmentation Patients Excluded from our RIAT Reanalysis and Reason for Exclusion.

|  | Bupropion SR | Buspirone |
| --- | --- | --- |
| **Number of patients included in NEJM analysis** | 279 | 286 |
| **Patients who should have been excluded:** | 63 | 61 |
| **Exclusion criteria #1: Patients with a baseline HRSD score <14 or missing a HRSD score at entry into step-1** | 38 | 45 |
| **Exclusion criteria #2: Patients who had an HRSD score <8 at entry into step-2** | 30 | 24 |
| **\*Number of patients included in RIAT Reanalysis** | 216 | 225 |

\* Bupropion SR had 5 patients and buspirone 8 who met both exclusion criteria.

Demographic and clinical characteristics of the 441 patients are described in Supplement 6. Similar to what the STAR*D investigators reported, we found a significant difference in mean symptom severity for medication-augmentation patients on the HRSD at entry into step-2 (*M*_1_=17.6; SD_1_=6.3) compared to the medication-switch patients (*M*_2_=20.3; SD_2_=6.2; *p* < 0.001).

### Step-2 medication-augmentation treatment

Table 2 presents the outcomes for remission, response, and mean change.

**Table 2:** Remission, Response, and Mean Change in Augmentation Patients.

|  | Bupropion SR plus Citalopram (N=216) |  | Buspirone plus Citalopram (N=225) |  | All Augmentation Patients(N=441) |  |
| --- | --- | --- | --- | --- | --- | --- |
|  | N | % | N | % | N | % |
| **HRSD Remission** | 54 | 25.0% | 58 | 25.8% | 112 | 25.4% |
| **HRSD Remission with Imputation\*** | 63 | 29.2% | 68 | 30.2% | 116 | 29.7% |
| **HRSD Response with Imputation\*** | 66 | 30.6% | 66 | 29.3% | 132 | 29.9% |
|  | Mean | SD | Mean | SD |  |  |
| **HRSD Mean Change with Imputation\*** | 4.3 | 7.4 | 4.4 | 6.9 |  |  |

\* For patients missing an exit HRSD score, their last QIDS-SR score is mapped to the HRSD and used to calculate the HRSD Remission with Imputation Rates, HRSD Response with Imputation Rates, and HRSD Mean Change with Imputation.

Our RIAT reanalysis using the protocol-defined remission criteria found that 25.0% of the 216 patients receiving bupropion SR remitted versus 25.8% of the 225 patients receiving buspirone. This difference was not statistically significant (χ^2^(1, *N*=441) = 0.04, *p* = 0.851).

Similarly, we found no significant differences on the sensitivity analysis with QIDS-SR-imputed missing HRSD scores for remission (χ^2^ [1, *N*=441] = 0.06, *p* = 0.808), response (χ^2^ [1, *N*=441] = 0.08, *p* = 0.779), or mean symptom change (*t*(434) = -0.15, *p* = 0.883).

We found numerically lower remission rates for the HRSD (primary outcome) in our RIAT reanalysis compared to those reported by the STAR*D investigators, but the differences were not significant for bupropion SR (29.7% vs 25.0%; χ^2^ [1, *N* = 495] = 1.39, *p* = 0.238) nor buspirone (30.1% vs 25.8%; χ^2^ [1, *N* = 511] = 1.15, *p* = 0.284). However, we did find a significantly lower remission rate compared to the NEJM publication on the secondary outcome measure (imputed HDRS scores) for bupropion SR (39.0% vs 29.2%; χ^2^ [1, *N* = 495] = 4.90, *p* = 0.027) but not for buspirone (32.9% vs 30.2%; χ^2^ [1, *N* = 511] = 0.41, *p* = 0.524).

Figure 1 compares the NEJM publication and our RIAT reanalysis first on the primary outcome measure for remission (the HRSD) and then the secondary measure (QIDS-SR for the NEJM publication and HRSD with imputation in our reanalysis).

### TESI rates for step-2 augmentation patients

Table 3 presents the TESI rates for the augmentation treatments.

**Table 3:** TESI Rates for Augmentation Treatments.

|  | **Bupropion SR**<br>**plus Citalopram** | **Buspirone plus**<br>**Citalopram** |
| --- | --- | --- |
| **Number of evaluable patients** | 216 | 225 |
| **Number of evaluable patients with a baseline QIDS-C suicide ideation score of 0 or 1. These patients are classified as non-suicidal at baseline.** | 168 | 173 |
| **Number of non-suicidal patients at baseline with one or more post-baseline QIDS-C suicide ideation scores of 2 or 3. This is the number of TESI patients.** | 6 | 24 |
| **TESI Rate** | 3.6% | 13.9% |

Our RIAT reanalysis found that the proportion of step-2 augmentation patients meeting criteria for TESI was significantly higher in the buspirone augmentation condition than the bupropion augmentation condition (13.9% vs 3.6%; (χ^2^(1, *N*=341) = 11.27, *p* = 0.001).

### Follow-up phase: Relapse and sustained remission

Table 4 presents the relapse and sustained remission rates during follow-up.

**Table 4:**
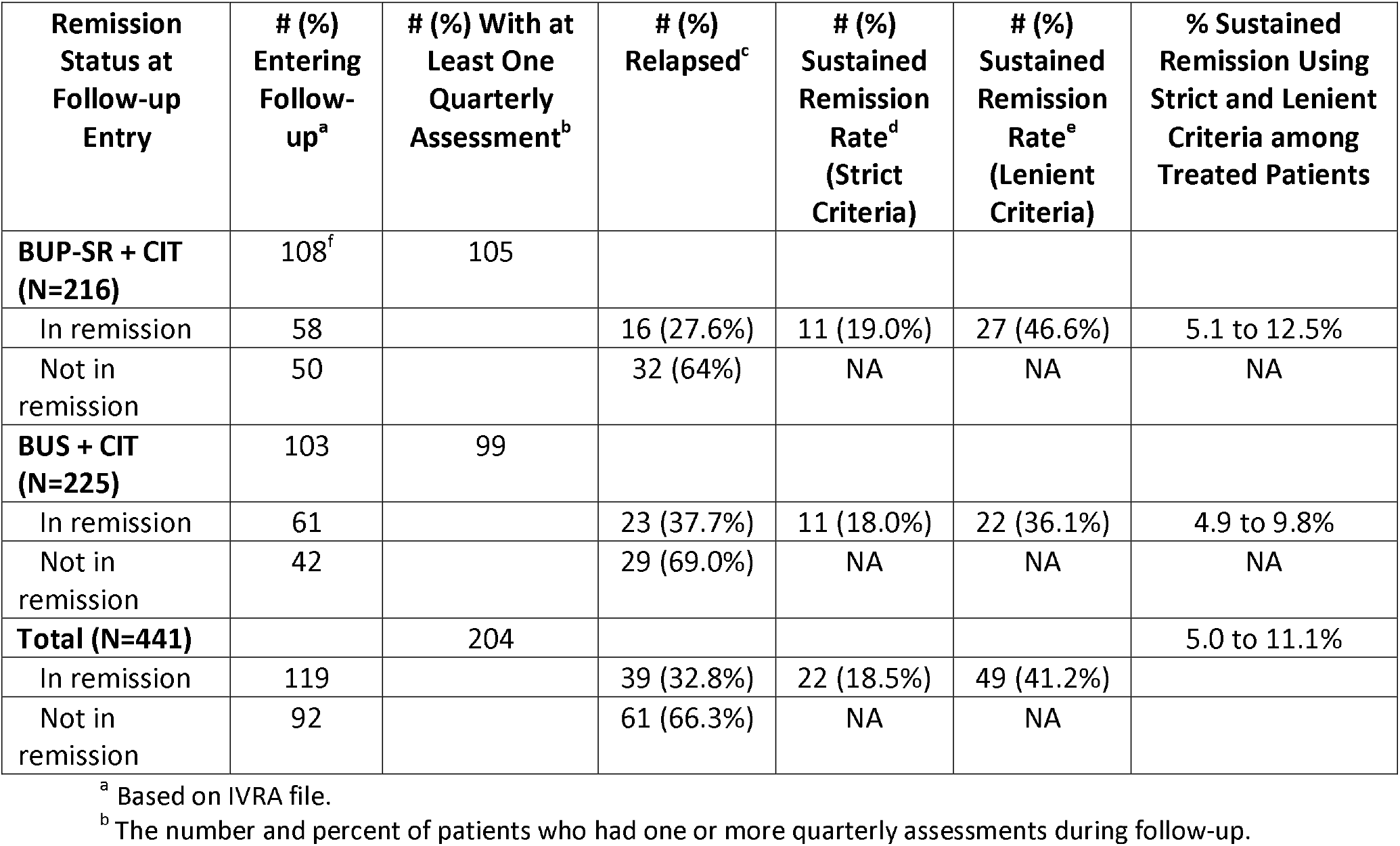
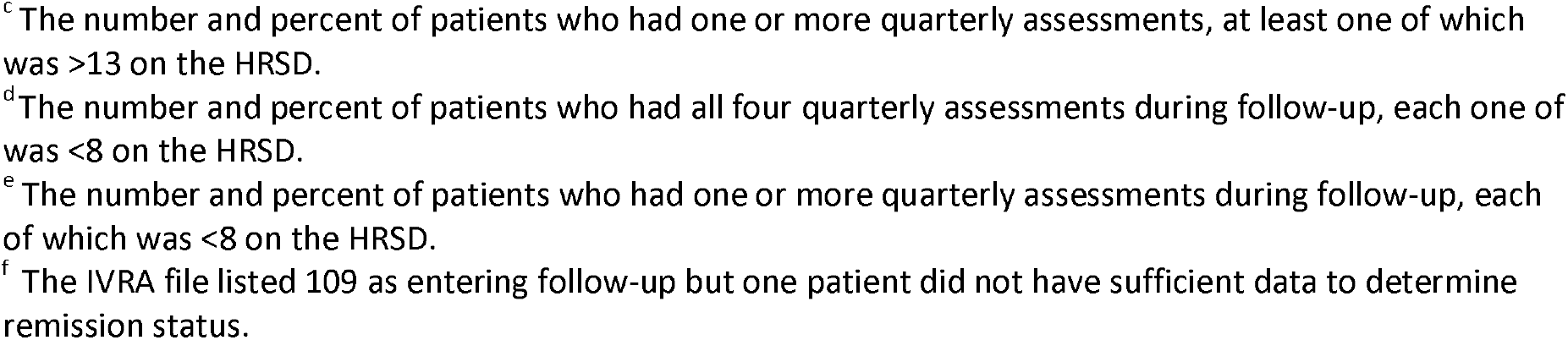
Relapse and Sustained Remission Rates During Follow-up.

| **Remission Status at Follow-up Entry** | **# (%) Entering Follow-up<sup>a</sup>** | **# (%) With at Least One Quarterly Assessment<sup>b</sup>** | **# (%) Relapsed<sup>c</sup>** | **# (%) Sustained Remission Rate<sup>d</sup> (Strict Criteria)** | **# (%) Sustained Remission Rate<sup>e</sup> (Lenient Criteria)** | **% Sustained Remission Using Strict and Lenient Criteria among Treated Patients** |
| --- | --- | --- | --- | --- | --- | --- |
| **BUP-SR + CIT (N=216)** | 108<sup>f</sup> | 105 |  |  |  |  |
| In remission | 58 |  | 16 (27.6%) | 11 (19.0%) | 27 (46.6%) | 5.1 to 12.5% |
| Not in remission | 50 |  | 32 (64%) | NA | NA | NA |
| **BUS + CIT (N=225)** | 103 | 99 |  |  |  |  |
| In remission | 61 |  | 23 (37.7%) | 11 (18.0%) | 22 (36.1%) | 4.9 to 9.8% |
| Not in remission | 42 |  | 29 (69.0%) | NA | NA | NA |
| **Total (N=441)** |  | 204 |  |  |  | 5.0 to 11.1% |
| In remission | 119 |  | 39 (32.8%) | 22 (18.5%) | 49 (41.2%) |  |
| Not in remission | 92 |  | 61 (66.3%) | NA | NA |  |

<sup>a</sup> Based on IVRA file.
<sup>b</sup> The number and percent of patients who had one or more quarterly assessments during follow-up.
<sup>c</sup> The number and percent of patients who had one or more quarterly assessments, at least one of which was >13 on the HRSD.
<sup>d</sup> The number and percent of patients who had all four quarterly assessments during follow-up, each one of which was <8 on the HRSD.
<sup>e</sup> The number and percent of patients who had one or more quarterly assessments during follow-up, each of which was <8 on the HRSD.
<sup>f</sup> The IVRA file listed 109 as entering follow-up but one patient did not have sufficient data to determine remission status.

Overall, 32.8% of patients in the two augmentation groups met remission criteria at entry into follow-up and relapsed (n=39). There was no significant difference in relapse rates between the bupropion and buspirone arms (χ^2^ [1, *N* = 119] = 1.38, *p* = 0.240).

There was also no significant difference in the proportions of patients who achieved a sustained remission between the augmentation arms when using the stringent criteria (χ^2^ [1, *N* = 119] = 0.02, *p* = 0.896) nor when using the lenient criteria (χ^2^ [1, *N* = 119] = 1.35, *p* = 0.245). Overall, of the 441 augmentation patients, the sustained remission rate during follow-up ranged from 5.0 to 11.1%, depending on the criteria used.

Of the 92 patients who did not meet remission criteria in step-2 augmentation treatment, 61 (66.3%) relapsed during the follow-up phase and there was no significant difference between the treatment groups (χ^2^ [1, *N* = 92] = 0.26, *p* = 0.608). However, the proportion of relapses was significantly greater among non-remitted than remitted patients (66.3% vs 32.8%; χ^2^ [1, *N* = 211] = 23.40, *p* < 0.001)

## Discussion

Our RIAT reanalysis of STAR*D’s step-2 augmentation patient-level dataset identified several important discrepancies from the original STAR*D published report.^3^

This includes 124 augmentation patients (21.9% of enrolled patients) that our RIAT reanalysis excluded because these patients failed to meet STAR*D’s inclusion for data analysis criteria, ^(see:1, figure 1)^ 54 of whom (9.6% of enrolled patients) scored as in remission on the HRSD prior to starting augmentation treatment despite the STAR*D investigators prespecifying that “patients who begin a level with HRSD <8 will be excluded from analyses” ^19, p.130^ (see Table 1).

Using STAR*D’s inclusion for data analysis criteria and protocol-specified primary outcome measure for remission (the HRSD), we found numerically lower rates for the step-2 augmentation treatments compared to those reported by the STAR*D investigators on the HRSD, albeit not a statistically significant difference.^3^ However, this difference was starker compared to the STAR*D summary papers which deviated from the protocol-specified inclusion criteria and used the QIDS-SR as outcome, without disclosing that this self-reported assessment was specifically excluded from being used to assess outcomes in the study protocol due to its role in guiding treatment decisions.^7,21^ In these seminal papers, remission rates were 39.0% for bupropion SR augmentation and 32.9% for buspirone augmentation, versus 25.0% and 25.8%, respectively, in this RIAT reanalysis with fidelity to the study protocol. Thus, for bupropion SR, the published QIDS-SR remission rate is 14 percentage points higher than the protocol-defined estimate (a 56% relative increase) and for buspirone, it is 7.1 percentage points higher (a 27% relative increase).

Furthermore, when using the protocol-specified blinded HRSD rather than the patient self-reported QIDS-SR, we found no significant difference in symptom change between bupropion SR and buspirone. This contrasts with the STAR*D investigators’ report of significantly greater symptom reduction associated with bupropion based on the QIDS-SR.

This reanalysis also found that the percentage of patients who both remitted during step-2 augmentation treatment and sustained the remission during follow-up ranged from only 4.9-12.5%, depending on the stringency of the criteria used—again, with no significant difference between treatments.

An unexpected finding was the significant difference in TESI rates between bupropion SR (3.6%) and buspirone (13.9%) when used to augment citalopram. The fourfold difference in TESI rates is important information in clinical decision making; particularly when there were no significant differences on any other clinically-relevant outcome (i.e., remission, response, symptom change, relapse, and sustained remission). We are not aware of potential explanations or other research findings of this significant difference; future studies should attempt to replicate and discern this difference.

It is unfortunate that despite having published TESI analyses of step-1 citalopram treatment,^15,22-24^ the STAR*D investigators did not report TESI rates in their steps 2-4 comparative effectiveness studies.^2-6^ Instead, the STAR*D investigators reported no significant differences on any clinically-relevant outcome in these studies and state that:

> *“****DEPRESSION*** *can be treated successfully by primary care physicians under “real world” conditions. Furthermore, the particular drug or drugs used are not as important as following a rational plan: giving antidepressant medications in adequate doses, monitoring the patient’s symptoms and side effects and adjusting the regimen accordingly, and switching drugs or adding new drugs to the regimen only after an adequate trial*.*”*^25, p.57, emphasis in the original^

Contrary to this conclusion and recommendations, our RIAT augmentation reanalysis suggests that what “*new drugs*” are added do in fact matter—*at least in terms of their impact on the likelihood of TESI*—as did our medication-switch reanalysis, which found a 43% relative increase in TESI rates when switching from citalopram to another antidepressant.^9^ We believe this is important information in the decision-making process for physicians and patients following a failed response to SSRI therapy.

A major limitation in the STAR*D trial is that there was no placebo-control group in any of the four treatment steps. Therefore, it is impossible to exclude spontaneous remission, the nonspecific effects of treatment, regression to the mean, or the extended use of citalopram alone as the likely explanation for these findings. As Kennedy et al. reported in their review, augmenting an antidepressant with bupropion or buspirone do not appear to outperform augmentation with placebo in double-blind RCTs, suggesting that the benefit seen in STAR*D may primarily reflect non-pharmacological factors like spontaneous improvement and placebo effects.^26^

In conclusion, this RIAT reanalysis of STAR*D step-2 augmentation treatments revealed lower remission rates than originally reported due to methodological deviations from the prespecified protocol by the STAR*D investigators. Moreover, we found no differences in effectiveness between treatments, with patients in both groups having a low likelihood of achieving a sustained remission during follow-up care. We did, however, find a clinically important difference in TESI rates between bupropion SR and buspirone augmentation which physicians and patients may want to incorporate in their decision-making process for those who decide to pursue medication augmentation following a failed trial on an SSRI.

The current findings add to our previous re-analyses which found that many of the original published findings of STAR*D deviate from those obtained with fidelity to the protocol. This, together with our findings for STAR*D’s TESI and long-term results, as well as findings from placebo-controlled RCTs, lead to different conclusions and recommendations than those put forward by the STAR*D investigators.

Given the world-wide impact of the STAR*D trial in treating MDD, the current RIAT reanalysis continues the process of restoring, with fidelity to the original study protocol, the actual STAR*D results for use in evidence-based treatment. We encourage other independent investigators to examine the STAR*D patient-level dataset as well.

## Data Availability

Data used in the preparation of this manuscript were downloaded in January 2025 from the controlled access datasets distributed from the NIMH-supported National Database for Clinical Trials (NDCT).

https://www.nimh.nih.gov/funding/clinical-research/researchers

https://www.nimh.nih.gov/funding/clinical-research/researchers

## Acknowledgements

Data used in the preparation of this manuscript were downloaded in January 2025 from the controlled access datasets distributed from the NIMH-supported National Database for Clinical Trials (NDCT). NDCT is a collaborative informatics system created by the National Institute of Mental Health to provide a national resource to support and accelerate discovery related to clinical trial research in mental health. The content of this publication does not necessarily reflect the views of the RIAT Support Center, NIMH, nor the National Institutes of Health.

## Contributors

HEP, JDA, CX, TK, MP, and IK contributed to the design of the study. CX and TK conducted all of the data analyses. HEP wrote the manuscript with input from KK, CX, MP, JDA, TK, and IK. HEP is responsible for the overall content as the guarantor.

