## Supplementary Appendix for "Restoring STAR*D: A RIAT Reanalysis of Medication Augmentation Therapy After Failed SSRI Treatment Using Patient-Level Data with Fidelity to the Original Research Protocol"

**Table of Contents**

1. Supplement 1:  Patient flow chart moving from step-1 citalopram treatment to the step-2 medication-augmentation therapies.
2. Supplement 2: STAR*D's system of care designed to maximize remissions while minimizing elapse and dropouts.
3. Supplement 3: Description of the medication-augmentation medications and their dosing.
4. Supplement 4: Medication-augmentation patients missing an exit HRSD.
5. Supplement 5: The statistical code used to analyze the STAR*D patient-level dataset.
6. Supplement 6: Demographic and clinical characteristics table.

### Supplement 1: Patient Flow Chart

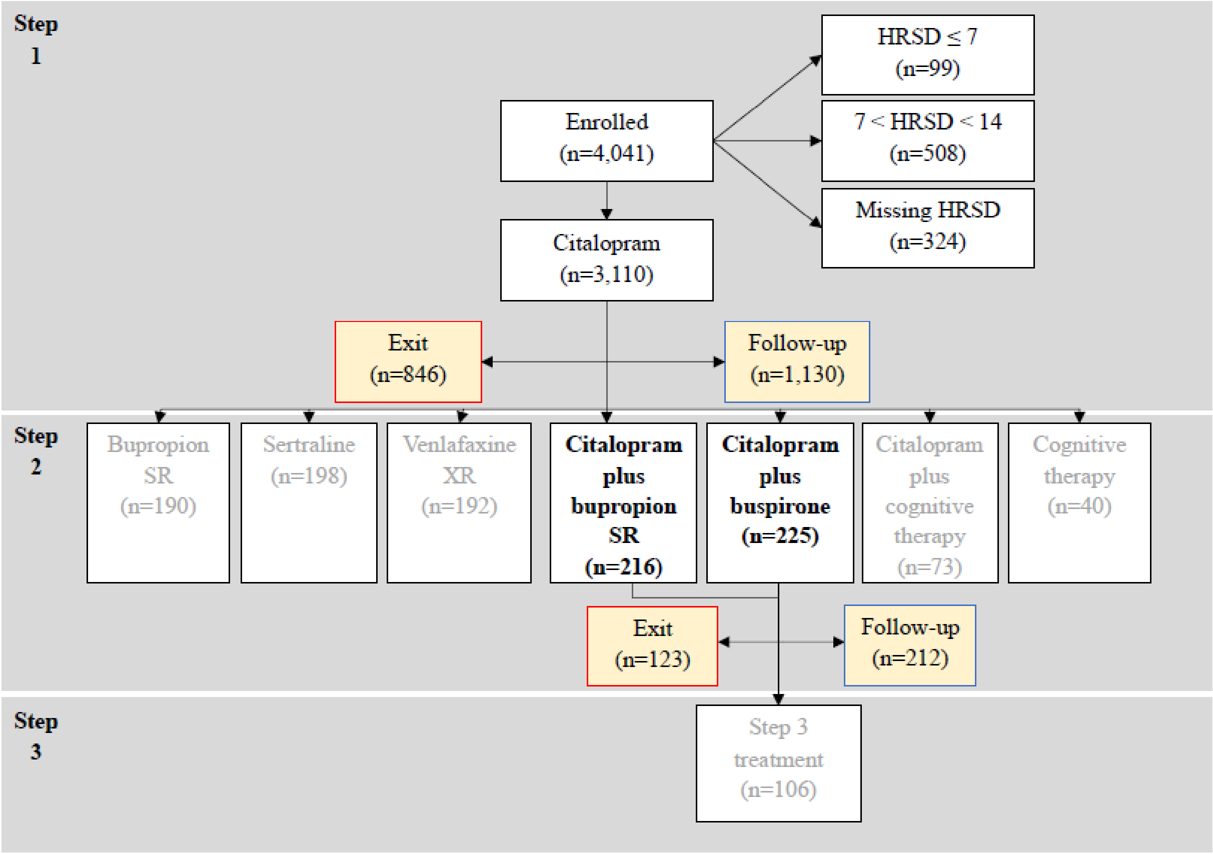

**Supplement 2: Highest Quality of Acute and Continuing-Care to Maximize Remissions While Minimizing Relapse and Dropouts**

| Descriptor | Explanation |
|---|---|
| Optimized Sustained Study Participation to Minimize Dropouts | • Promoted patients' study affiliation via STAR*D-branded brochures, bimonthly newsletters, and an informational video emphasizing STAR*D's public health significance and the critical role played by patients;<br>• Provided a multistep educational program for patients and families throughout acute-care based on the neurochemical imbalance theory of depression that included "*a glossy visual representation of the brain and neurotransmitters*," consistently emphasizing that "*depression is a disease, like diabetes or high blood pressure, and has not been caused by something the patient has or has not done. (Depression is an illness, not a personal weakness or character flaw*.) *The educator should emphasize that depression can be treated as effectively as other illnesses*," and "*explaining the basic principles of mechanism of action*" for the patient's current antidepressant drug (Patient Education Manual pp. 4–7).;<br>• Used a letter reminder system to alert patients before appointments in those clinics without such systems who had a >15% rate of missed appointments;<br>• Ensured timely follow-up and rescheduling of missed appointments by calling patients on the day of the missed appointment, and again within 24 hours, if there was no response. Patient's physician sent letter within 48 hours if contact was not established;<br>• Used a letter reminder system for all research outcome assessment calls during acute and continuing-care;<br>• In every clinic visit, the Clinical Research Coordinator (CRC) discussed the research outcomes phone calls with the patient to ensure that the calls were completed on schedule and worked to resolve any problematic issues regarding said calls [Clinical Procedures Manual, page 75];<br>• Paid patients $25.00 for participating in each telephonic research outcomes assessment;<br>• Permitted patients to re-enter acute and/or continuing-care within four weeks after having dropped out [Clinical Procedures Manual, page 80]; |

| | |
|---|---|
| | • Recommended one-year of continuing-care for all patients who achieved a satisfactory clinical response with the essential goal of preventing relapse [Clinical Procedures Manual, page 15] and<br>• Permitted continuing-care patients to remain in the study if they moved from the area [Clinical Procedures Manual, page 81]. |
| Acute-Care Visits | Physicians met with patients on entry into each new step to initiate drug treatment with follow-up visits scheduled on weeks 2, 4, 6, 9, 12, with an optional week 14 visit. |
| Measurement-Based Care | Conducted structured evaluations of symptoms and side-effects at each visit and included a centralized treatment monitoring and physician feedback system to ensure consistent implementation of optimal care across research sites. |
| Aggressive Medication Dosing | Provided aggressive medication dosing with a fully adequate dose for a sufficient duration to "*ensure that the likelihood of achieving remission was maximized and that those who did not reach remission were truly resistant to the medication*". |
| Liberal Prescribing of Non-Study Medications | Physicians had great leeway in prescribing non-study medications to treat comorbid symptoms resulting in:<br>• 22.7% taking Trazodone for sleep;<br>• 12.2% taking an anti-anxiety medication;<br>• 18.1% taking either a sedative or hypnotic medication; and<br>• An undisclosed percent taking medications to address side-effects. |
| Continuing-Care Visits | Patients saw their physician every 2 months and continued taking their treatment medication(s) at the same doses but their physicians were allowed to make any psychotherapy, medication, and/or medication dose changes to maximize the likelihood of maintaining patients' remission status. Additional continuing-care visits were scheduled when patients began to experience a return of depressive symptoms and/or intolerable side-effects [Clinical Procedures Manual. page 78]. |
| Clinical Research Coordinator (CRC) | Each site had a CRC who:<br>• Saw patients before each visit administering multiple measures to them including the QIDS-SR during each acute-care visit;<br>• Assisted physicians in protocol implementation; and<br>• Provided patients support and encouragement in protocol implementation. |
| Treatment Designed to Enhance Subject Retention | Treatment was designed to minimize drop-outs and/or non-compliance including:<br>• Open label prescribing during acute and continuing-care with no placebo control condition during any study phase; |

| | |
|---|---|
| | • Patients chose their acceptable treatment assignments for steps two and three to eliminate any concerns they might have about receiving an unacceptable assignment.<br>• During each step, patients could enroll immediately into the next step if they had intolerable side-effects or had maximized their current medication(s)' dosing without achieving a remission; and<br>• During any step, patients could enter continuing-care directly on their current medication(s) if they were treatment responders even if they had not achieved remission. This was done to minimize responders from dropping out in order to avoid having to discontinue their current medication(s) and start a new drug regimen. |

O'Neal, B., & Biggs, M. (2001). STAR*D patient education manual. Retrieved July 5, 2010, from http://www.edc.pitt.edu/stard/public/study_manuals.html

Trivedi MH, Stegman D, Rush AJ, Wisniewski SR, Nierenberg AA: STAR*D clinical procedures manual. July 31, 2002. www.edc.pitt. edu/stard/public/study_manuals.html

Trazodone 68 (24.5) 60 (21.1)
Anxiolytics 40 (14.4) 29 (10.2)
Sedatives or hypnotics 51 (18.4) 51 (17.9)

### Supplement 3:
### Description of Medication-Augmentation Treatments

**Step-2 Citalopram augmentation treatments**:

During the augmentation trial, the citalopram dose was kept constant but reduced if side effects developed. The level 2 augmentation treatments were:

- Buspirone (Buspar), a partial agonist at the postsynaptic 5-hydroxytryptamine 1A (5-HT1A) receptor that is believed to enhance the activity of SSRIs through the 5HT1A receptors. The starting dose was 15 mg per day week 1, increasing to 30 mg per day week 2, and then to 45 mg per day for weeks 3 through 5, and a final, maximum dose of 60 mg per day week 6 and onward.
- Sustained-release bupropion (Wellbutrin SR) whose neurochemical action mechanisms are unknown but is believed to produce antidepressant effects by blocking the reuptake of dopamine and norepinephrine. The initial dose was 200 mg per day during weeks 1 and 2, increasing to 300 mg per day by week 4 and to 400 mg per day week 6 and onward.

**Supplement 4: Number and Percent of Medication-Augmentation Patients Missing Exit HRSD**

| | #/(%) with Missing Exit HRSD |
|---|---|
| Bupropion SR (N=216) | 58 (26.9%) |
| Buspirone (N=225) | 52 (23.1%) |
| Total (N=441) | 110 (24.9%) |

**Supplement 5:**
**Definitions of Acute and Follow-up Care Outcomes and the Statistical Code Used to Analyze the Dataset**

Response was defined as a ≥ 50% reduction in HRSD between baseline level 2 HRSD (defined as the exit HRSD from level 1, or if they are missing exit HRSD from level, the HRSD mapped from QIDS at the last observation of level 1) and the exit for level 2 HRSD (defined as the last HRSD for level 2, or if they are missing HRSD from that level, the HRSD mapped from the QIDS at the last observation of level 2).

We also calculated mean change in HRSD score between baseline level 2 HRSD (defined as the exit HRSD from the previous level, in this case, level 1, or if they are missing exit HRSD from level, the HRSD mapped from QIDS at the last observation of the previous level) and the exit for level 2 HRSD (defined as the last HRSD for level 2, or if they are missing HRSD from that level, the HRSD mapped from the QIDS at the last observation of level 2).

```
##################################################################################################
###

####################                  READING IN RAW HRSD DATA
       ####################

##################################################################################################
###



setwd("STARD Package_1235145/")



### read in p01 version hrsd Data

hrsdNewHeader         <-      read.table("hrsd01.txt", nrows = 1, header = FALSE,
stringsAsFactors = FALSE)             #Extract current file header

hrsdNewRaw                   <-      read.table("hrsd01.txt", skip = 2, header = FALSE, fill =
T)                                           #Extract current file data

colnames(hrsdNewRaw )  <- unlist(hrsdNewHeader)
                       #Merge file Fheader with data


### read in ivra data

ivraNewHeader         <-      read.table("ivra01.txt", nrows = 1, header = FALSE,
stringsAsFactors = FALSE)             #Extract current file header

ivraNewRaw                   <-      read.table("ivra01.txt", skip = 2, header = FALSE, fill =
T)                                           #Extract current file data

colnames(ivraNewRaw )  <- unlist(ivraNewHeader)
                       #Merge file header with data
```

```
### read in QIDS data

qidsNewHeader          <-      read.table("qids01.txt", nrows = 1, header = FALSE,
stringsAsFactors = FALSE)             #Extract current file header

qidsNewRaw                    <-      read.table("qids01.txt", skip = 2, header = FALSE, fill =
T)                                          #Extract current file data

colnames(qidsNewRaw )  <- unlist(qidsNewHeader)
                       #Merge file header with data









##########################################################################################

####################          TASK 1: RESCORING VARIABLES           ###################

##########################################################################################


##################  SCORING HRSD-17          ################

# Since hdtot_r variable is missing for all levels except enrollment, we need to manually rescore
HRSD

#



## Find out how many have hdtot_r

table(is.na(hrsdNewRaw$hdtot_r))                     # Turns out only 4039 scored hdtot_r's
available


## Find which levels have legitimate hdtot_r ##

table(hrsdNewRaw[!is.na(hrsdNewRaw$hdtot_r),"level"])               # turns out only enrollment
has the HRSD sum score



## Look only at enrollment data since they have a valid hdtot_r     ##

table(hrsdNewRaw[hrsdNewRaw$level =="Enrollment",]$hdtot_r , useNA="always")
```

```
## Rescoring by adding symptom vars   ##

hrsdSymptomVars        <- c("hsoin", "hmnin", "hemin", "hmdsd", "hpanx", "hinsg", "happt", "hwl",
"hsanx", "hhypc", "hvwsf", "hsuic", "hintr", "hengy", "hslow", "hagit", "hsex")

hrsdNewRaw$hrsdtot_sum_NAasNA                <- rowSums(hrsdNewRaw[,c(hrsdSymptomVars)])
        #This version treats NA's as zeros

### Checking IVRA -- Action_call shows when people are moved to followup OR to new treatment
        ###

with(ivraNewRaw, table(action_call, txassign))

##################  Scoring QIDS data since Followup is lacking qstot
        ################

# Since followup QIDs is laking a qstot totalscore, we need to manually score QIDS data

#

# Sleep variables

qidsNewRaw $maxSleep   <- apply(qidsNewRaw [, c("vsoin", "vmnin", "vhysm", "vemin")], MARGIN = 1,
max, na.rm = T)

# Add that result to the maximum score of the weight/appetite variables (SAPDC, SAPIN, SWTDC,
SWTIN)

qidsNewRaw $maxAppetite       <- apply(qidsNewRaw [, c("vapdc", "vapin", "vwtdc", "vwtin")],
MARGIN = 1, max, na.rm = T)

# Then add that sum to the maximum of the two motor variables (SAGIT, SSLOW)

qidsNewRaw $maxMotor   <- apply(qidsNewRaw [, c("vagit", "vslow")], MARGIN = 1, max, na.rm = T)

# And finally add to that sum the sum of the rest of the scores (SMDSD, SCNTR, SVWSF, SSUIC,
SINTR, SENGY)

qidsNewRaw $sumOther   <- rowSums(qidsNewRaw [, c("vmdsd", "vcntr", "vvwsf", "vsuic", "vintr",
"vengy")])

qidsNewRaw $SR_TOTAL <- rowSums(qidsNewRaw [, c("maxSleep", "maxAppetite", "maxMotor",
"sumOther")])
```

```
######################################################################
##############        Function to remap QIDS        ##################
######################################################################

# In order to impute missing HRSD scores using QIDS scores, as per methods section of BMJ Open paper

remapHRSD_QIDS <- function(x){
       y <- NA
       x <- as.character(x)
       switch(x,
              '1' =  {y <- 1.5},
              '2' =  {y <- 3},
              '3' =  {y <- 4},
              '4' =  {y <- 5.5},
              '5' =  {y <- 7},
              '6' =  {y <- 8},
              '7' =  {y <- 9.5},
              '8' =  {y <- 11},
              '9' =  {y <- 12},
              '10' =  {y <- 13},
              '11' =  {y <- 14.5},
              '12' =  {y <- 16},
              '13' =  {y <- 17},
              '14' =  {y <- 18.5},
              '15' =  {y <- 18.5},
              '16' =  {y <- 20},
              '17' =  {y <- 21.5},
              '18' =  {y <- 23},
              '19' =  {y <- 24},
              '20' =  {y <- 25},
```

```
            '21' =  {y <- 26.5},
            '22' =  {y <- 28},
            '23' =  {y <- 29},
            '24' =  {y <- 30.5},
            '25' =  {y <- 32},
            '26' =  {y <- 34},
            '27' =  {y <- 44})
        return(y)
        }




##################################################################################################
#########
##################            DEFINE WINDOWS FOR FOLLOW UP HRSD
        #####################################
##################################################################################################
#########



###     Define month windows for QIDS ###
#
# Follow up HRSD and QIDS
# Find out if they have a month corresponding HRSD. If they do not, then replace it with a QIDS
from the specified window (SR_TOTAL). (Have as two outcomes)
#
# 4 time points:
# Month 3 - Window 63-119
# Month 6 - Window 154-210
# Month 9 - Window 245-301
# Month 12 - Window 336-392
#
# If they have observation not in windows, count it as missing
#
# For month on hrsd variable, use month variable (which describes monthin follow up, -2 for those
who do not have followup)
# For days, use days_baseline from QIDS
#
```

```
######################################

       qidsWindowMonth3_lo    <- 63
       qidsWindowMonth3_hi    <- 119
       qidsWindowMonth6_lo    <- 154
       qidsWindowMonth6_hi    <- 210
       qidsWindowMonth9_lo    <- 245
       qidsWindowMonth9_hi    <- 301
       qidsWindowMonth12_lo   <- 336
       qidsWindowMonth12_hi   <- 392

       qidsWindowMonth3_mid   <- 91
       qidsWindowMonth6_mid   <- 182
       qidsWindowMonth9_mid   <- 273
       qidsWindowMonth12_mid  <- 364




#################################################################################################
##########################
####################          TASK 2: Pulling out HRSD scores, calculating remissions
               ###################
#################################################################################################
###########################

# First define all subjectID's
allHRSD_subjectID <- unique(hrsdNewRaw$src_subject_id)


# Create an output file for HRSD

stardOutputs   <- data.frame(src_subject_id = allHRSD_subjectID,

                                                    meetInclusion  = NA,

                                                    # IVRA level entry variables
```

```
maxlevel_ivra= NA,

level1_ivra_days_enter = NA,
level2_ivra_days_enter = NA,
level2A_ivra_days_enter = NA,
level3_ivra_days_enter = NA,
level4_ivra_days_enter = NA,
fu_ivra_days_enter   = NA,

level1_ivra_entered = NA,
level2_ivra_entered = NA,
level2A_ivra_entered = NA,
level3_ivra_entered = NA,
level4_ivra_entered = NA,
fu_ivra_entered  = NA,

# Level 1 variables


level1_first_hrsd_day = NA,
level1_first_hrsd_sum = NA,
level1_last_hrsd_day = NA,
level1_last_hrsd_sum = NA,

level1_nHRSD   = NA,



level1_last_qids_day   = NA,
level1_last_qids_qstot = NA,

level1_tx = NA,
level1_remitted = NA,

level1_remitted_imputed = NA,
level1_first_hrsd_imputedQIDS = NA,
level1_last_hrsd_imputedQIDS = NA,
```

```
level1_responded = NA,
level1_hrsdchange = NA,

level1_responded_imputed = NA,
level1_hrsdchange_imputed = NA,


# Level 2 variables


level2_hasHRSDData = NA,
level2_last_hrsd_day = NA,
level2_last_hrsd_sum= NA,
level2_last_qids_day   = NA,
level2_last_qids_qstot = NA,

level2_tx = NA,
level2_remitted = NA,



level2_remitted_imputed = NA,
level2_last_hrsd_imputedQIDS = NA,
level2_responded = NA,
level2_hrsdchange = NA,

level2_responded_imputed = NA,
level2_hrsdchange_imputed = NA,



# Level 2A variables


level2A_hasHRSDData = NA,
level2A_last_hrsd_day = NA,
level2A_last_hrsd_sum= NA,
level2A_last_qids_day  = NA,
level2A_last_qids_qstot = NA,
```

```
level2A_tx = NA,
level2A_remitted = NA,

level2A_remitted_imputed = NA,
level2A_last_hrsd_imputedQIDS = NA,
level2A_responded = NA,
level2A_hrsdchange = NA,

level2A_responded_imputed = NA,
level2A_hrsdchange_imputed = NA,


# Level 3 variables

level3_hasHRSDData = NA,
level3_last_hrsd_day = NA,
level3_last_hrsd_sum= NA,

level3_last_qids_day   = NA,
level3_last_qids_qstot = NA,

level3_tx = NA,
level3_remitted = NA,

level3_remitted_imputed = NA,
level3_last_hrsd_imputedQIDS = NA,
level3_responded = NA,
level3_hrsdchange = NA,

level3_responded_imputed = NA,
level3_hrsdchange_imputed = NA,


# Level 4 variables

level4_hasHRSDData = NA,
level4_last_hrsd_day = NA,
```

```
level4_last_hrsd_sum= NA,
level4_last_qids_day   = NA,
level4_last_qids_qstot = NA,

level4_tx = NA,
level4_remitted = NA,

level4_remitted_imputed = NA,
level4_last_hrsd_imputedQIDS = NA,

level4_responded = NA,
level4_hrsdchange = NA,

level4_responded_imputed = NA,
level4_hrsdchange_imputed = NA,

# Followup

fu_hasHRSDData = NA,
fu_hrsd_month3 = NA,
fu_hrsd_month6 = NA,
fu_hrsd_month9 = NA,
fu_hrsd_month12 = NA,
fu_qids_month3 = NA,
fu_qids_month6 = NA,
fu_qids_month9 = NA,
fu_qids_month12 = NA,
fu_qids_month3_date = NA,
fu_qids_month6_date = NA,
fu_qids_month9_date = NA,
fu_qids_month12_date = NA,

fu_noObservations = NA,

fu_hrsdImputed_month3 = NA,
fu_hrsdImputed_month6 = NA,
fu_hrsdImputed_month9 = NA,
```

```
                                                      fu_hrsdImputed_month12 = NA,



                                                      fu_remissionbeforeFU   = NA,
                                                      fu_remissionbeforeFU_imputed = NA,

                                                      fu_relapse = NA,
                                                      fu_susRemission_full = NA,
                                                      fu_susRemission_weak = NA,

                                                      flagged = NA)




# Now loop through subjectID's in Level1     #
for(i in 1:length(allHRSD_subjectID )){


        # Pull out current subject ID
        currentID                           <- allHRSD_subjectID[i]


        #Pull out Level1 data, for current subject


        currentSubjectHRSD_all        <- hrsdNewRaw[hrsdNewRaw$src_subject_id == currentID,]


        currentSubjectHRSD_Level1            <- hrsdNewRaw[hrsdNewRaw$level == "Level 1" &
hrsdNewRaw$src_subject_id == currentID,]
        currentSubjectHRSD_Level2            <- hrsdNewRaw[hrsdNewRaw$level == "Level 2" &
hrsdNewRaw$src_subject_id == currentID,]
        currentSubjectHRSD_Level2A    <- hrsdNewRaw[hrsdNewRaw$level == "Level 2A" &
hrsdNewRaw$src_subject_id == currentID,]
        currentSubjectHRSD_Level3            <- hrsdNewRaw[hrsdNewRaw$level == "Level 3" &
hrsdNewRaw$src_subject_id == currentID,]
        currentSubjectHRSD_Level4            <- hrsdNewRaw[hrsdNewRaw$level == "Level 4" &
hrsdNewRaw$src_subject_id == currentID,]


        currentSubjectHRSD_fu  <- hrsdNewRaw[hrsdNewRaw$level == "Follow up" &
hrsdNewRaw$src_subject_id == currentID,]
```

```
##########     IVRA LEVELS            ##########



#Pull out the ivra data for participant

currentSubject_ivra           <- ivraNewRaw[ivraNewRaw$src_subject_id == currentID,]


#Pull out corresponding day someone entered a level

level1_ivra_days_enter  <-  unique(currentSubject_ivra[currentSubject_ivra$level == "Level 1" & currentSubject_ivra$action_call == 1, "days_baseline"])

level2_ivra_days_enter  <-  unique(currentSubject_ivra[currentSubject_ivra$level == "Level 2" & currentSubject_ivra$action_call == 1 , "days_baseline"])

level2A_ivra_days_enter <-  unique(currentSubject_ivra[currentSubject_ivra$level == "Level 2A" & currentSubject_ivra$action_call == 1, "days_baseline"])

level3_ivra_days_enter  <-  unique(currentSubject_ivra[currentSubject_ivra$level == "Level 3" & currentSubject_ivra$action_call == 1, "days_baseline"])

level4_ivra_days_enter  <-  unique(currentSubject_ivra[currentSubject_ivra$level == "Level 4" & currentSubject_ivra$action_call == 1, "days_baseline"])

fu_ivra_days_enter  <-  unique(currentSubject_ivra[currentSubject_ivra$level == "Follow up" & currentSubject_ivra$action_call == 2, "days_baseline"])




# Clean up IVRA for people who did not enter a level

level1_ivra_days_enter   <- ifelse(length(level1_ivra_days_enter) != 0, level1_ivra_days_enter , NA)

level2_ivra_days_enter   <- ifelse(length(level2_ivra_days_enter) != 0, level2_ivra_days_enter , NA)

level2A_ivra_days_enter   <- ifelse(length(level2A_ivra_days_enter) != 0, level2A_ivra_days_enter , NA)

level3_ivra_days_enter   <- ifelse(length(level3_ivra_days_enter) != 0, level3_ivra_days_enter , NA)

level4_ivra_days_enter   <- ifelse(length(level4_ivra_days_enter) != 0, level4_ivra_days_enter , NA)

fu_ivra_days_enter    <- ifelse(length(level4_ivra_days_enter) != 0, fu_ivra_days_enter , NA)



# binary to specify if they entered a level or not

level1_ivra_entered  <- ifelse(nrow(currentSubject_ivra[currentSubject_ivra$action_call == 1 & currentSubject_ivra$level == "Level 1",]) > 0, 1,0)

level2_ivra_entered  <- ifelse(nrow(currentSubject_ivra[currentSubject_ivra$action_call == 1 & currentSubject_ivra$level == "Level 2",]) > 0, 1,0)
```

```
        level2A_ivra_entered  <- ifelse(nrow(currentSubject_ivra[currentSubject_ivra$action_call
== 1 & currentSubject_ivra$level == "Level 2A",]) > 0, 1,0)

        level3_ivra_entered  <- ifelse(nrow(currentSubject_ivra[currentSubject_ivra$action_call
== 1 & currentSubject_ivra$level == "Level 3",]) > 0, 1,0)

        level4_ivra_entered  <- ifelse(nrow(currentSubject_ivra[currentSubject_ivra$action_call
== 1 & currentSubject_ivra$level == "Level 4",]) > 0, 1,0)

        fu_ivra_entered               <-
ifelse(nrow(currentSubject_ivra[currentSubject_ivra$action_call == 2 & currentSubject_ivra$level
== "Follow up",]) > 0, 1,0)


        #Calculate max level, we work backyards incase someone has skipped a level

        maxlevel_ivra          <- ifelse(level4_ivra_entered == 1 , "Level 4",

                                      ifelse(level3_ivra_entered == 1 , "Level 3",

                                      ifelse(level2A_ivra_entered == 1 , "Level 2A",

                                      ifelse(level2_ivra_entered == 1 , "Level 2",

                                      ifelse(level1_ivra_entered == 1 , "Level 1", NA)))))




        ### save IVRA outputs  ###

        stardOutputs[stardOutputs$src_subject_id == currentID, "maxlevel_ivra"] <- maxlevel_ivra


        stardOutputs[stardOutputs$src_subject_id == currentID, "level1_ivra_days_enter"]   <-
level1_ivra_days_enter

        stardOutputs[stardOutputs$src_subject_id == currentID, "level2_ivra_days_enter"]   <-
level2_ivra_days_enter

        stardOutputs[stardOutputs$src_subject_id == currentID, "level2A_ivra_days_enter"]  <-
level2A_ivra_days_enter

        stardOutputs[stardOutputs$src_subject_id == currentID, "level3_ivra_days_enter"]   <-
level3_ivra_days_enter

        stardOutputs[stardOutputs$src_subject_id == currentID, "level4_ivra_days_enter"]   <-
level4_ivra_days_enter

        stardOutputs[stardOutputs$src_subject_id == currentID, "fu_ivra_days_enter"]
        <- fu_ivra_days_enter



        stardOutputs[stardOutputs$src_subject_id == currentID, "level1_ivra_entered"] <-
level1_ivra_entered

        stardOutputs[stardOutputs$src_subject_id == currentID, "level2_ivra_entered"] <-
level2_ivra_entered

        stardOutputs[stardOutputs$src_subject_id == currentID, "level2A_ivra_entered"] <-
level2A_ivra_entered
```

```
        stardOutputs[stardOutputs$src_subject_id == currentID, "level3_ivra_entered"] <-
level3_ivra_entered

        stardOutputs[stardOutputs$src_subject_id == currentID, "level4_ivra_entered"] <-
level4_ivra_entered

        stardOutputs[stardOutputs$src_subject_id == currentID, "fu_ivra_entered"]  <-
fu_ivra_entered



        ##########      HRSD DATA            ##########


        ### HRSD Level 1 ###


        #Count how many HRSD Rows in level 1

        level1_nHRSD   <- nrow(unique(currentSubjectHRSD_Level1))


        #Select only rows where days_baseline <= 5

        currentSubjectHRSD_L1_firstLevel15   <-
unique(currentSubjectHRSD_Level1[currentSubjectHRSD_Level1$days_baseline <=5 ,])


        #For subjects with multiple before days_baseline <=5, collapse both dates so we can see

        level1_multiple_daysbaseline_5days   <-
paste(currentSubjectHRSD_L1_firstLevel15$days_baseline, collapse = "-")

        level1_multiple_hrsd_5days           <-
paste(currentSubjectHRSD_L1_firstLevel15$hrsdtot_sum_NAasNA, collapse = "-")



        #We want the earliest one, so pick the earliest row (if applicable)

        currentSubject_L1_earliestHRSD       <-
        unique(currentSubjectHRSD_L1_firstLevel15[order(currentSubjectHRSD_L1_firstLevel15$days_b
aseline),])[1,]


        level1_first_hrsd_day <- currentSubject_L1_earliestHRSD$days_baseline

        level1_first_hrsd_sum <- currentSubject_L1_earliestHRSD$hrsdtot_sum_NAasNA




        #Find the latest HRSD day

        level1_latestHRSD_day  <- unique(max(currentSubjectHRSD_Level1$days_baseline))
```

```
        # Pull out row matching lastLevel1 day, make sure it is only unique. If multple rows then
pull out the firstLevel1.

        level1_last_hrsd_rows  <-
unique(currentSubjectHRSD_Level1[currentSubjectHRSD_Level1$days_baseline ==
        level1_latestHRSD_day & !is.na(currentSubjectHRSD_Level1$days_baseline ) &
!is.na(currentSubjectHRSD_Level1$hrsdtot_sum_NAasNA),])[1,]


        # Require lastLevel1 HRSD to be over 10 days

        level1_last_hrsd_rows  <- level1_last_hrsd_rows[level1_last_hrsd_rows$days_baseline > 10,]


        if (nrow(level1_last_hrsd_rows) == 0 ){

                level1_last_hrsd_day          <- NA

                level1_last_hrsd_sum   <- NA

        }else {

                level1_last_hrsd_day                  <- level1_last_hrsd_rows$days_baseline

                level1_last_hrsd_sum                  <- level1_last_hrsd_rows$hrsdtot_sum_NAasNA

        }




        # Check if they meet inclusion criteria

        meetInclusion  <-      ifelse(is.na(currentSubject_L1_earliestHRSD$hrsdtot_sum_NAasNA) |
currentSubject_L1_earliestHRSD$hrsdtot_sum_NAasNA <  14,0,1)


        # Check if they remitted in level 1

        level1_remitted <- ifelse(meetInclusion == 1 & !is.na(level1_last_hrsd_sum) &
level1_last_hrsd_sum<= 7, 1,

                                        ifelse(is.na(level1_last_hrsd_sum), NA, 0))

        # Find level 1 treatment condition

        level1_tx <- ivraNewRaw[ivraNewRaw$src_subject_id == currentID & ivraNewRaw$level ==
"Level 1" & ivraNewRaw$txassign != "", "txassign"]


        # Calculate HRSD change

        level1_hrsdchange      <- level1_last_hrsd_sum - level1_first_hrsd_sum


        # Calculate response, defined as 50% improvement from last to first

        level1_responded       <- ifelse((-level1_hrsdchange / level1_first_hrsd_sum) >= 0.50, 1,
0)


        #Now save this to HRSD Output File
```

```
        stardOutputs[stardOutputs$src_subject_id == currentID, "level1_first_hrsd_day"]
        <- level1_first_hrsd_day

        stardOutputs[stardOutputs$src_subject_id == currentID, "level1_first_hrsd_sum"]
        <- level1_first_hrsd_sum


        stardOutputs[stardOutputs$src_subject_id == currentID, "level1_last_hrsd_sum"]
        <- level1_last_hrsd_sum

        stardOutputs[stardOutputs$src_subject_id == currentID, "level1_last_hrsd_day"]
        <-  level1_last_hrsd_day

        stardOutputs[stardOutputs$src_subject_id == currentID, "level1_nHRSD"]
        <- level1_nHRSD

        stardOutputs[stardOutputs$src_subject_id == currentID, "level1_remitted"]
        <- level1_remitted


        stardOutputs[stardOutputs$src_subject_id == currentID, "level1_hrsdchange"]
        <- level1_hrsdchange

        stardOutputs[stardOutputs$src_subject_id == currentID, "level1_responded"]
        <- level1_responded


        stardOutputs[stardOutputs$src_subject_id == currentID, "meetInclusion"]
        <- meetInclusion

        stardOutputs[stardOutputs$src_subject_id == currentID, "level1_tx"]
        <- level1_tx



        ### Now do Level 2 HRSD ###



        if(nrow(currentSubjectHRSD_Level2) == 0){            # If they do not have level 2,
specify everything as NA


                level2_hasHRSDData     <- 0

                level2_last_hrsd_sum   <-  NA

                level2_last_hrsd_day   <-  NA

                level2_remitted               <-  NA

                level2_responded              <- NA

                level2_hrsdchange             <- NA


        }else {                #Otherwise calculate level 2 data


                level2_hasHRSDData     <- 1
```

```
            #Find the latest HRSD day

            level2_last_hrsd_day   <- unique(max(currentSubjectHRSD_Level2$days_baseline))


            # Pull out row matching last day, make sure it is only unique. If multple rows
then pull out the first.

            level2_last_hrsd_rows  <-
unique(currentSubjectHRSD_Level2[currentSubjectHRSD_Level2$days_baseline == level2_last_hrsd_day&
!is.na(currentSubjectHRSD_Level2$days_baseline ) ,])[1,]




            # Pull out last level 2 hrsd sum

            level2_last_hrsd_sum<- level2_last_hrsd_rows$ hrsdtot_sum_NAasNA


            ##Calculate level 2 remitted: meetInclusion = 1, did not remit in level 1, and
level 2 last HRSD <= 7

            level2_remitted <- ifelse(meetInclusion == 1  & level2_last_hrsd_sum<= 7, 1, 0)


            # Calculate HRSD change in level 2 -- using last level 1 HRSD as level 2 baseline

            level2_hrsdchange      <- level2_last_hrsd_sum - level1_last_hrsd_sum


            # Calculate response, defined as 50% improvement from last level 1 HRSD

            level2_responded       <- ifelse((-level2_hrsdchange / level1_last_hrsd_sum) >=
0.50, 1, 0)


        }


        # Pull out level2 treatment

        level2_tx <- ivraNewRaw[ivraNewRaw$src_subject_id == currentID & ivraNewRaw$level ==
"Level 2" & ivraNewRaw$txassign != "", "txassign"]


        # Convert level 2 treatment to NA for people who did not get assigned

        level2_tx <- ifelse(length(level2_tx) != 0, level2_tx, NA)




        ## Now saving it to output

        stardOutputs[stardOutputs$src_subject_id == currentID, "level2_hasHRSDData"]
        <- level2_hasHRSDData
```

```
        stardOutputs[stardOutputs$src_subject_id == currentID, "level2_last_hrsd_sum"]
        <- level2_last_hrsd_sum

        stardOutputs[stardOutputs$src_subject_id == currentID, "level2_last_hrsd_day"]
        <- level2_last_hrsd_day

        stardOutputs[stardOutputs$src_subject_id == currentID, "level2_remitted"]
        <- level2_remitted

        stardOutputs[stardOutputs$src_subject_id == currentID, "level2_tx"]
                <- level2_tx


        stardOutputs[stardOutputs$src_subject_id == currentID, "level2_hrsdchange"]
                <- level2_hrsdchange

        stardOutputs[stardOutputs$src_subject_id == currentID, "level2_responded"]
                <- level2_responded



        ### Now do Level 2 A HRSD ###



        if(nrow(currentSubjectHRSD_Level2A) == 0){          # If they do not have Level 2 A,
specify everything as NA

                level2A_hasHRSDData   <- 0

                level2A_last_hrsd_sum  <-  NA

                level2A_last_hrsd_day  <-  NA

                level2A_remitted              <-  NA


                level2A_responded             <- NA

                level2A_hrsdchange            <- NA


        }else {                #Otherwise calculate Level 2 A data


                level2A_hasHRSDData   <- 1




                #Find the latest HRSD day

                level2A_last_hrsd_day  <- unique(max(currentSubjectHRSD_Level2A$days_baseline))


                # Pull out row matching last day, make sure it is only unique. If multple rows
then pull out the first.

                level2A_last_hrsd_rows <-
unique(currentSubjectHRSD_Level2A[currentSubjectHRSD_Level2A$days_baseline ==
level2A_last_hrsd_day& !is.na(currentSubjectHRSD_Level2A$days_baseline ) ,])[1,]
```

```
            # Pull out last Level 2 A hrsd sum
            level2A_last_hrsd_sum <- level2A_last_hrsd_rows$ hrsdtot_sum_NAasNA


            ##Calculate Level 2 A remitted: meetInclusion = 1, did not remit in level 1, and
Level 2 A last HRSD <= 7
            level2A_remitted <- ifelse(meetInclusion == 1 & level2A_last_hrsd_sum <= 7, 1, 0)


            # Calculate HRSD change from last level 2 HRSD
            level2A_hrsdchange     <- level2A_last_hrsd_sum - level2_last_hrsd_sum


            # Calculate response, defined as 50% improvement from last level 2 HRSD
            level2A_responded      <- ifelse((-level2A_hrsdchange / level2_last_hrsd_sum) >=
0.50, 1, 0)



        }


        # Pull out level2A treatment
        level2A_tx <- ivraNewRaw[ivraNewRaw$src_subject_id == currentID & ivraNewRaw$level ==
"Level 2A" & ivraNewRaw$txassign != "", "txassign"]


        # Convert Level 2 A treatment to NA for people who did not get assigned
        level2A_tx <- ifelse(length(level2A_tx) != 0, level2A_tx, NA)



        ## Now saving it to output
        stardOutputs[stardOutputs$src_subject_id == currentID, "level2A_hasHRSDData"]
        <- level2A_hasHRSDData
        stardOutputs[stardOutputs$src_subject_id == currentID, "level2A_last_hrsd_sum"]
        <- level2A_last_hrsd_sum
        stardOutputs[stardOutputs$src_subject_id == currentID, "level2A_last_hrsd_day"]
        <- level2A_last_hrsd_day
        stardOutputs[stardOutputs$src_subject_id == currentID, "level2A_remitted"]
        <- level2A_remitted
        stardOutputs[stardOutputs$src_subject_id == currentID, "level2A_tx"]
        <- level2A_tx


        stardOutputs[stardOutputs$src_subject_id == currentID, "level2A_hrsdchange"]
        <- level2A_hrsdchange
```

```
        stardOutputs[stardOutputs$src_subject_id == currentID, "level2A_responded"]
        <- level2A_responded



        ### Now do Level 3 HRSD ###



        if(nrow(currentSubjectHRSD_Level3) == 0){            # If they do not have Level 3,
specify everything as NA

                level3_hasHRSDData     <- 0

                level3_last_hrsd_sum   <-  NA

                level3_last_hrsd_day   <-  NA

                level3_remitted               <-  NA

                level3_hrsdchange             <-  NA

                level3_responded              <-  NA

                level3_entry_hrsd_sum  <-  NA


        }else {                #Otherwise calculate Level 3 data


                level3_hasHRSDData     <- 1



                #Find the latest HRSD day

                level3_last_hrsd_day   <- unique(max(currentSubjectHRSD_Level3$days_baseline))


                # Pull out row matching last day, make sure it is only unique. If multple rows
then pull out the first.

                level3_last_hrsd_rows  <-
unique(currentSubjectHRSD_Level3[currentSubjectHRSD_Level3$days_baseline == level3_last_hrsd_day&
!is.na(currentSubjectHRSD_Level3$days_baseline ) ,])[1,]



                # Pull out last Level 3 hrsd sum

                level3_last_hrsd_sum <- level3_last_hrsd_rows$ hrsdtot_sum_NAasNA


                ##Calculate Level 3 remitted: meetInclusion = 1, did not remit in level 1, and
Level 3 last HRSD <= 7

                level3_remitted <- ifelse(meetInclusion == 1 & level3_last_hrsd_sum <= 7, 1, 0)
```

```
                ## Find level 3 entry HRSD depending on if they entered level 2A or not

                level3_entry_hrsd_sum  <- ifelse(level2A_hasHRSDData == 1, level2A_last_hrsd_sum ,
level2_last_hrsd_sum )


                # Calculate HRSD change

                level3_hrsdchange      <- level3_last_hrsd_sum - level3_entry_hrsd_sum


                # Calculate response, defined as 50% improvement from last to first

                level3_responded       <- ifelse((-level3_hrsdchange / level3_entry_hrsd_sum ) >=
0.50, 1, 0)



        }


        # Pull out level3 treatment

        level3_tx <- ivraNewRaw[ivraNewRaw$src_subject_id == currentID & ivraNewRaw$level ==
"Level 3" & ivraNewRaw$txassign != "", "txassign"]


        # Convert Level 3 treatment to NA for people who did not get assigned

        level3_tx <- ifelse(length(level3_tx) != 0, level3_tx, NA)



        ## Now saving it to output

        stardOutputs[stardOutputs$src_subject_id == currentID, "level3_hasHRSDData"]
                <- level3_hasHRSDData

        stardOutputs[stardOutputs$src_subject_id == currentID, "level3_last_hrsd_sum"]
                <- level3_last_hrsd_sum

        stardOutputs[stardOutputs$src_subject_id == currentID, "level3_last_hrsd_day"]
                <- level3_last_hrsd_day

        stardOutputs[stardOutputs$src_subject_id == currentID, "level3_remitted"]
        <- level3_remitted

        stardOutputs[stardOutputs$src_subject_id == currentID, "level3_tx"]
                <- level3_tx


        stardOutputs[stardOutputs$src_subject_id == currentID, "level3_hrsdchange"]
                <- level3_hrsdchange

        stardOutputs[stardOutputs$src_subject_id == currentID, "level3_responded"]
                <- level3_responded


        ### Now do Level 4 HRSD ###
```

```
        if(nrow(currentSubjectHRSD_Level4) == 0){            # If they do not have Level 4, 
specify everything as NA

                level4_hasHRSDData     <- 0

                level4_last_hrsd_sum   <-  NA

                level4_last_hrsd_day   <-  NA

                level4_remitted                <-  NA

                level4_hrsdchange              <-  NA

                level4_responded               <-  NA


        }else {                #Otherwise calculate Level 4 data


                level4_hasHRSDData     <- 1




                #Find the latest HRSD day

                level4_last_hrsd_day   <- unique(max(currentSubjectHRSD_Level4$days_baseline))


                # Pull out row matching last day, make sure it is only unique. If multple rows 
then pull out the first.

                level4_last_hrsd_rows  <- 
unique(currentSubjectHRSD_Level4[currentSubjectHRSD_Level4$days_baseline == level4_last_hrsd_day& 
!is.na(currentSubjectHRSD_Level4$days_baseline ) ,])[1,]




                # Pull out last Level 4 hrsd sum

                level4_last_hrsd_sum <- level4_last_hrsd_rows$ hrsdtot_sum_NAasNA


                ##Calculate Level 4 remitted: meetInclusion = 1, did not remit in level 1, and 
Level 4 last HRSD <= 7

                level4_remitted <- ifelse(meetInclusion == 1 & level4_last_hrsd_sum <= 7, 1, 0)


                # Calculate HRSD change

                level4_hrsdchange      <- level4_last_hrsd_sum - level3_last_hrsd_sum


                # Calculate response, defined as 50% improvement from last to first

                level4_responded       <- ifelse((-level4_hrsdchange / level3_last_hrsd_sum) >= 
0.50, 1, 0)
```

```
        }

        # Pull out level4 treatment

        level4_tx <- ivraNewRaw[ivraNewRaw$src_subject_id == currentID & ivraNewRaw$level ==
"Level 4" & ivraNewRaw$txassign != "", "txassign"]

        # Convert Level 4 treatment to NA for people who did not get assigned

        level4_tx <- ifelse(length(level4_tx) != 0, level4_tx, NA)

        ## Now saving it to output

        stardOutputs[stardOutputs$src_subject_id == currentID, "level4_hasHRSDData"]
                <- level4_hasHRSDData

        stardOutputs[stardOutputs$src_subject_id == currentID, "level4_last_hrsd_sum"]
        <- level4_last_hrsd_sum

        stardOutputs[stardOutputs$src_subject_id == currentID, "level4_last_hrsd_day"]
        <- level4_last_hrsd_day

        stardOutputs[stardOutputs$src_subject_id == currentID, "level4_remitted"]
        <- level4_remitted

        stardOutputs[stardOutputs$src_subject_id == currentID, "level4_tx"]
                <- level4_tx

        stardOutputs[stardOutputs$src_subject_id == currentID, "level4_hrsdchange"]
                <- level4_hrsdchange

        stardOutputs[stardOutputs$src_subject_id == currentID, "level4_responded"]
                        <- level4_responded

        ########       Adding QIDS Data              ########

        #Pull out qids rows for coresponding subject, but only self report

        currentSubjectQids     <- qidsNewRaw[qidsNewRaw$version_form == "Self Rating" &
qidsNewRaw$src_subject_id == currentID & !is.na(qidsNewRaw$qstot),]

                if(level1_ivra_entered ==1 ){                       ##Test if they have entered
level 1 according to IVRA

                        level1_last_qids_day   <- max(currentSubjectQids[currentSubjectQids$level
== "Level 1", "days_baseline"])

                        level1_last_qids_qstot <- currentSubjectQids[currentSubjectQids$level ==
"Level 1" & currentSubjectQids$days_baseline == level1_last_qids_day ,"qstot"][1]
```

```
                        stardOutputs[stardOutputs$src_subject_id ==
currentID,"level1_last_qids_day"]     <- level1_last_qids_day

                        stardOutputs[stardOutputs$src_subject_id ==
currentID,"level1_last_qids_qstot"] <- level1_last_qids_qstot




                        #Pull date of firstqids

                        level1_first_qids_day  <-
unique(min(currentSubjectQids[currentSubjectQids$level == "Level 1", "days_baseline"]))

                        level1_first_qids_qstot       <-
currentSubjectQids[currentSubjectQids$level == "Level 1" & currentSubjectQids$days_baseline ==
level1_first_qids_day ,"qstot"][1]




                        #If they are missing a last HRSD, impute it using the QIDS

                        level1_last_hrsd_imputedQIDS  <- ifelse(!is.na(level1_last_hrsd_sum),
level1_last_hrsd_sum, remapHRSD_QIDS(level1_last_qids_qstot))

                        level1_first_hrsd_sum_imputed <- ifelse(!is.na(level1_first_hrsd_sum),
level1_first_hrsd_sum, remapHRSD_QIDS(level1_first_qids_qstot))


                        level1_remitted_imputed <- ifelse(meetInclusion == 1 &
!is.na(level1_last_hrsd_imputedQIDS  ) & level1_last_hrsd_imputedQIDS  <= 7, 1,
        ifelse(is.na(level1_last_hrsd_imputedQIDS  ), NA, 0))


                        level1_hrsdchange_imputed     <- level1_last_hrsd_imputedQIDS   -
level1_first_hrsd_sum_imputed

                        level1_responded_imputed      <- ifelse((-level1_hrsdchange_imputed /
level1_first_hrsd_sum_imputed) >= 0.50, 1, 0)




                        stardOutputs[stardOutputs$src_subject_id ==
currentID,"level1_last_hrsd_imputedQIDS"]           <- level1_last_hrsd_imputedQIDS

                        stardOutputs[stardOutputs$src_subject_id ==
currentID,"level1_remitted_imputed"]         <- level1_remitted_imputed

                        stardOutputs[stardOutputs$src_subject_id ==
currentID,"level1_hrsdchange_imputed"]              <- level1_hrsdchange_imputed

                        stardOutputs[stardOutputs$src_subject_id ==
currentID,"level1_responded_imputed"]               <- level1_responded_imputed






                }else {

                        stardOutputs[stardOutputs$src_subject_id ==
currentID,"level1_last_qids_day"]                   <- NA
```

```
                    stardOutputs[stardOutputs$src_subject_id ==
currentID,"level1_last_qids_qstot"]          <- NA
                    stardOutputs[stardOutputs$src_subject_id ==
currentID,"level1_last_hrsd_imputedQIDS"]           <- NA
                    stardOutputs[stardOutputs$src_subject_id ==
currentID,"level1_remitted_imputed"]         <- NA


                    stardOutputs[stardOutputs$src_subject_id ==
currentID,"level1_hrsdchange_imputed"]              <- NA
                    stardOutputs[stardOutputs$src_subject_id ==
currentID,"level1_responded_imputed"]               <- NA



               }


               if(level2_ivra_entered ==1 ){
                    level2_last_qids_day   <- max(currentSubjectQids[currentSubjectQids$level
== "Level 2", "days_baseline"])
                    level2_last_qids_qstot <- currentSubjectQids[currentSubjectQids$level ==
"Level 2" & currentSubjectQids$days_baseline == level2_last_qids_day ,"qstot"][1]
                    stardOutputs[stardOutputs$src_subject_id ==
currentID,"level2_last_qids_day"]     <- level2_last_qids_day
                    stardOutputs[stardOutputs$src_subject_id ==
currentID,"level2_last_qids_qstot"] <- level2_last_qids_qstot


                    level2_last_hrsd_imputedQIDS  <- ifelse(!is.na(level2_last_hrsd_sum),
level2_last_hrsd_sum, remapHRSD_QIDS(level2_last_qids_qstot))
                    level2_remitted_imputed <- ifelse(meetInclusion == 1 &
level2_last_hrsd_imputedQIDS  <= 7, 1,       ifelse(is.na(level2_last_hrsd_imputedQIDS  ), NA,
0))


                    level2_hrsdchange_imputed     <- level2_last_hrsd_imputedQIDS   -
level1_last_hrsd_imputedQIDS
                    level2_responded_imputed      <- ifelse((-level2_hrsdchange_imputed /
level1_last_hrsd_imputedQIDS   ) >= 0.50, 1, 0)


                    stardOutputs[stardOutputs$src_subject_id ==
currentID,"level2_last_hrsd_imputedQIDS"]           <- level2_last_hrsd_imputedQIDS
                    stardOutputs[stardOutputs$src_subject_id ==
currentID,"level2_remitted_imputed"]         <- level2_remitted_imputed


                    stardOutputs[stardOutputs$src_subject_id ==
currentID,"level2_hrsdchange_imputed"]              <- level2_hrsdchange_imputed
                    stardOutputs[stardOutputs$src_subject_id ==
currentID,"level2_responded_imputed"]               <- level2_responded_imputed
```

```
                }else {

                        stardOutputs[stardOutputs$src_subject_id ==
currentID,"level2_last_qids_day"]     <- NA

                        stardOutputs[stardOutputs$src_subject_id ==
currentID,"level2_last_qids_qstot"]   <- NA

                        stardOutputs[stardOutputs$src_subject_id ==
currentID,"level2_last_hrsd_imputedQIDS"]    <- NA

                        stardOutputs[stardOutputs$src_subject_id ==
currentID,"level2_remitted_imputed"]         <- NA

                        stardOutputs[stardOutputs$src_subject_id ==
currentID,"level2_hrsdchange_imputed"]       <- NA

                        stardOutputs[stardOutputs$src_subject_id ==
currentID,"level2_responded_imputed"]                <- NA




                }

                if(level2A_ivra_entered ==1 ){

                        level2A_last_qids_day  <- max(currentSubjectQids[currentSubjectQids$level
== "Level 2 A", "days_baseline"])

                        level2A_last_qids_qstot       <-
currentSubjectQids[currentSubjectQids$level == "Level 2 A" & currentSubjectQids$days_baseline ==
level2A_last_qids_day ,"qstot"][1]

                        stardOutputs[stardOutputs$src_subject_id ==
currentID,"level2A_last_qids_day"]    <- level2A_last_qids_day

                        stardOutputs[stardOutputs$src_subject_id ==
currentID,"level2A_last_qids_qstot"] <- level2A_last_qids_qstot



                        level2A_last_hrsd_imputedQIDS  <- ifelse(!is.na(level2A_last_hrsd_sum),
level2A_last_hrsd_sum, remapHRSD_QIDS(level2A_last_qids_qstot))

                        level2A_remitted_imputed <- ifelse(meetInclusion == 1 &
level2A_last_hrsd_imputedQIDS  <= 7, 1, ifelse(is.na(level2A_last_hrsd_imputedQIDS  ), NA, 0))



                        level2A_hrsdchange_imputed           <- level2A_last_hrsd_imputedQIDS   -
level2_last_hrsd_imputedQIDS

                        level2A_responded_imputed            <- ifelse((-
level2A_hrsdchange_imputed / level2_last_hrsd_imputedQIDS   ) >= 0.50, 1, 0)




                        stardOutputs[stardOutputs$src_subject_id ==
currentID,"level2A_last_hrsd_imputedQIDS"]   <- level2A_last_hrsd_imputedQIDS

                        stardOutputs[stardOutputs$src_subject_id ==
currentID,"level2A_remitted_imputed"]                <- level2A_remitted_imputed
```

```
                        stardOutputs[stardOutputs$src_subject_id ==
currentID,"level2A_hrsdchange_imputed"]             <- level2A_hrsdchange_imputed

                        stardOutputs[stardOutputs$src_subject_id ==
currentID,"level2A_responded_imputed"]              <- level2A_responded_imputed



                }else {

                        stardOutputs[stardOutputs$src_subject_id ==
currentID,"level2A_last_qids_day"]    <- NA

                        stardOutputs[stardOutputs$src_subject_id ==
currentID,"level2A_last_qids_qstot"]  <- NA

                        stardOutputs[stardOutputs$src_subject_id ==
currentID,"level2A_last_hrsd_imputedQIDS"]   <- NA

                        stardOutputs[stardOutputs$src_subject_id ==
currentID,"level2A_remitted_imputed"]               <- NA


                        stardOutputs[stardOutputs$src_subject_id ==
currentID,"level2A_hrsdchange_imputed"]      <- NA

                        stardOutputs[stardOutputs$src_subject_id ==
currentID,"level2A_responded_imputed"]              <- NA



                }


                if(level3_ivra_entered ==1 ){

                        level3_last_qids_day   <- max(currentSubjectQids[currentSubjectQids$level
== "Level 3", "days_baseline"])

                        level3_last_qids_qstot <- currentSubjectQids[currentSubjectQids$level ==
"Level 3" & currentSubjectQids$days_baseline == level3_last_qids_day ,"qstot"][1]

                        stardOutputs[stardOutputs$src_subject_id ==
currentID,"level3_last_qids_day"]     <- level3_last_qids_day

                        stardOutputs[stardOutputs$src_subject_id ==
currentID,"level3_last_qids_qstot"] <- level3_last_qids_qstot


                        level3_last_hrsd_imputedQIDS  <- ifelse(!is.na(level3_last_hrsd_sum),
level3_last_hrsd_sum, remapHRSD_QIDS(level3_last_qids_qstot))

                        level3_remitted_imputed <- ifelse(meetInclusion == 1 &
level3_last_hrsd_imputedQIDS  <= 7, 1,       ifelse(is.na(level3_last_hrsd_imputedQIDS  ), NA,
0))


                        # Create level 3 entry variable depending on if they entered 2A or just 2

                        level3_entry_hrsd_sum_imputed <- ifelse(level2A_ivra_entered == 1,
level2A_last_hrsd_imputedQIDS, level2_last_hrsd_imputedQIDS)
```

```
                        level3_hrsdchange_imputed             <- level3_last_hrsd_imputedQIDS   -
level3_entry_hrsd_sum_imputed

                        level3_responded_imputed              <- ifelse((-level3_hrsdchange_imputed
/ level3_entry_hrsd_sum_imputed) >= 0.50, 1, 0)



                        stardOutputs[stardOutputs$src_subject_id ==
currentID,"level3_last_hrsd_imputedQIDS"]    <- level3_last_hrsd_imputedQIDS

                        stardOutputs[stardOutputs$src_subject_id ==
currentID,"level3_remitted_imputed"]         <- level3_remitted_imputed

                        stardOutputs[stardOutputs$src_subject_id ==
currentID,"level3_hrsdchange_imputed"]              <- level3_hrsdchange_imputed

                        stardOutputs[stardOutputs$src_subject_id ==
currentID,"level3_responded_imputed"]               <- level3_responded_imputed

               }else {

                        stardOutputs[stardOutputs$src_subject_id ==
currentID,"level3_last_qids_day"]     <- NA

                        stardOutputs[stardOutputs$src_subject_id ==
currentID,"level3_last_qids_qstot"]   <- NA

                        stardOutputs[stardOutputs$src_subject_id ==
currentID,"level3_last_hrsd_imputedQIDS"]    <- NA

                        stardOutputs[stardOutputs$src_subject_id ==
currentID,"level3_remitted_imputed"]         <- NA

                        stardOutputs[stardOutputs$src_subject_id ==
currentID,"level3_hrsdchange_imputed"]       <- NA

                        stardOutputs[stardOutputs$src_subject_id ==
currentID,"level3_responded_imputed"]               <- NA

               }

               if(level4_ivra_entered ==1 ){

                        level4_last_qids_day   <- max(currentSubjectQids[currentSubjectQids$level
== "Level 4", "days_baseline"])

                        level4_last_qids_qstot <- currentSubjectQids[currentSubjectQids$level ==
"Level 4" & currentSubjectQids$days_baseline == level4_last_qids_day ,"qstot"][1]

                        stardOutputs[stardOutputs$src_subject_id ==
currentID,"level4_last_qids_day"]     <- level4_last_qids_day

                        stardOutputs[stardOutputs$src_subject_id ==
currentID,"level4_last_qids_qstot"] <- level4_last_qids_qstot



                        level4_last_hrsd_imputedQIDS  <- ifelse(!is.na(level4_last_hrsd_sum),
level4_last_hrsd_sum, remapHRSD_QIDS(level4_last_qids_qstot))

                        level4_remitted_imputed <- ifelse(meetInclusion == 1 &
level4_last_hrsd_imputedQIDS  <= 7, 1,       ifelse(is.na(level4_last_hrsd_imputedQIDS  ), NA,
0))



                        level4_hrsdchange_imputed             <- level4_last_hrsd_imputedQIDS   -
level3_last_hrsd_imputedQIDS
```

```
			level4_responded_imputed		<- ifelse((-level4_hrsdchange_imputed / level3_last_hrsd_imputedQIDS   ) >= 0.50, 1, 0)



			stardOutputs[stardOutputs$src_subject_id == currentID,"level4_last_hrsd_imputedQIDS"]		<- level4_last_hrsd_imputedQIDS
			stardOutputs[stardOutputs$src_subject_id == currentID,"level4_remitted_imputed"]		<- level4_remitted_imputed
			stardOutputs[stardOutputs$src_subject_id == currentID,"level4_hrsdchange_imputed"]		<- level4_hrsdchange_imputed
			stardOutputs[stardOutputs$src_subject_id == currentID,"level4_responded_imputed"]		<- level4_responded_imputed


		}else {
			stardOutputs[stardOutputs$src_subject_id == currentID,"level4_last_qids_day"]	<- NA
			stardOutputs[stardOutputs$src_subject_id == currentID,"level4_last_qids_qstot"]	<- NA
			stardOutputs[stardOutputs$src_subject_id == currentID,"level4_last_hrsd_imputedQIDS"]	<- NA
			stardOutputs[stardOutputs$src_subject_id == currentID,"level4_remitted_imputed"]	<- NA
			stardOutputs[stardOutputs$src_subject_id == currentID,"level4_hrsdchange_imputed"]	<- NA
			stardOutputs[stardOutputs$src_subject_id == currentID,"level4_responded_imputed"]		<- NA
		}


		########	Followup Data  ########


		# Do they have follow HRSD Data recorded
		fu_hasHRSDData <- ifelse(nrow(currentSubjectHRSD_fu > 0) ,1 ,0)


		if(fu_ivra_entered  == 1){		#Check if they entered followup as recorded on IVRA


			currentSubjectQIDS_fu  <- qidsNewRaw[qidsNewRaw$src_subject_id == currentID& qidsNewRaw$level == "Follow-Up",]


			## Pull out matching followup HRSD for patient
			fu_hrsd_month3		<- currentSubjectHRSD_fu[currentSubjectHRSD_fu$month ==3 & !is.na(currentSubjectHRSD_fu$month),]$hrsdtot_sum_NAasNA
```

```
			fu_hrsd_month6		<- currentSubjectHRSD_fu[currentSubjectHRSD_fu$month
==6 & !is.na(currentSubjectHRSD_fu$month),]$hrsdtot_sum_NAasNA

			fu_hrsd_month9		<- currentSubjectHRSD_fu[currentSubjectHRSD_fu$month
==9 & !is.na(currentSubjectHRSD_fu$month),]$hrsdtot_sum_NAasNA

			fu_hrsd_month12		<-
currentSubjectHRSD_fu[currentSubjectHRSD_fu$month ==12 &
!is.na(currentSubjectHRSD_fu$month),]$hrsdtot_sum_NAasNA


			#Identify observations falling within qids windows


			fu_qids_month3_obs		<-
currentSubjectQIDS_fu[currentSubjectQIDS_fu$days_baseline >= qidsWindowMonth3_lo &
currentSubjectQIDS_fu$days_baseline  <=  qidsWindowMonth3_hi,]

			fu_qids_month6_obs		<-
currentSubjectQIDS_fu[currentSubjectQIDS_fu$days_baseline >= qidsWindowMonth6_lo &
currentSubjectQIDS_fu$days_baseline  <=  qidsWindowMonth6_hi,]

			fu_qids_month9_obs		<-
currentSubjectQIDS_fu[currentSubjectQIDS_fu$days_baseline >= qidsWindowMonth9_lo &
currentSubjectQIDS_fu$days_baseline  <=  qidsWindowMonth9_hi,]

			fu_qids_month12_obs		<-
currentSubjectQIDS_fu[currentSubjectQIDS_fu$days_baseline >= qidsWindowMonth12_lo &
currentSubjectQIDS_fu$days_baseline  <=  qidsWindowMonth12_hi,]


			fu_qids_month3_minDiff <- min(abs(fu_qids_month3_obs$days_baseline -
qidsWindowMonth3_mid))

			fu_qids_month6_minDiff <- min(abs(fu_qids_month6_obs$days_baseline -
qidsWindowMonth6_mid))

			fu_qids_month9_minDiff <- min(abs(fu_qids_month9_obs$days_baseline -
qidsWindowMonth9_mid))

			fu_qids_month12_minDiff		<- min(abs(fu_qids_month12_obs$days_baseline
- qidsWindowMonth12_mid))


			fu_qids_month3_index <- which(abs(fu_qids_month3_obs$days_baseline -
qidsWindowMonth3_mid) == fu_qids_month3_minDiff)

			fu_qids_month6_index <- which(abs(fu_qids_month6_obs$days_baseline -
qidsWindowMonth6_mid) == fu_qids_month6_minDiff)

			fu_qids_month9_index <- which(abs(fu_qids_month9_obs$days_baseline -
qidsWindowMonth9_mid) == fu_qids_month9_minDiff)

			fu_qids_month12_index <- which(abs(fu_qids_month12_obs$days_baseline -
qidsWindowMonth12_mid) == fu_qids_month12_minDiff)


			fu_qids_month3		<-	fu_qids_month3_obs[fu_qids_month3_index
,]$SR_TOTAL

			fu_qids_month3_date	<-	fu_qids_month3_obs[fu_qids_month3_index
,]$days_baseline

			fu_qids_month6		<-	fu_qids_month6_obs[fu_qids_month6_index
,]$SR_TOTAL
```

```
			fu_qids_month6_date	<-	fu_qids_month6_obs[fu_qids_month6_index
,]$days_baseline
			fu_qids_month9		<-	fu_qids_month9_obs[fu_qids_month9_index
,]$SR_TOTAL
			fu_qids_month9_date	<-	fu_qids_month9_obs[fu_qids_month9_index
,]$days_baseline
			fu_qids_month12			<-
	fu_qids_month12_obs[fu_qids_month12_index ,]$SR_TOTAL
			fu_qids_month12_date	<-	fu_qids_month12_obs[fu_qids_month12_index
,]$days_baseline


			# Removing null observations to NA	#


			fu_hrsd_month3 <- ifelse(is.null(fu_hrsd_month3), NA, fu_hrsd_month3)
			fu_hrsd_month6		<- ifelse(is.null(fu_hrsd_month6), NA,
fu_hrsd_month6)
			fu_hrsd_month9 <- ifelse(is.null(fu_hrsd_month9), NA, fu_hrsd_month9)
			fu_hrsd_month12		<- ifelse(is.null(fu_hrsd_month12), NA,
fu_hrsd_month12)


			fu_qids_month3 <- ifelse(is.null(fu_qids_month3), NA, fu_qids_month3)
			fu_qids_month6 <- ifelse(is.null(fu_qids_month6), NA, fu_qids_month6)
			fu_qids_month9 <- ifelse(is.null(fu_qids_month9), NA, fu_qids_month9)
			fu_qids_month12		<- ifelse(is.null(fu_qids_month12), NA,
fu_qids_month12)


			fu_qids_month3_date	<- ifelse(is.null(fu_qids_month3_date), NA,
fu_qids_month3_date)
			fu_qids_month6_date	<- ifelse(is.null(fu_qids_month6_date), NA,
fu_qids_month6_date)
			fu_qids_month9_date	<- ifelse(is.null(fu_qids_month9_date), NA,
fu_qids_month9_date)
			fu_qids_month12_date	<- ifelse(is.null(fu_qids_month12_date), NA,
fu_qids_month12_date)


			# Remapping qids to HRSD where missing HRSD
			fu_hrsdImputed_month3	<- ifelse(!is.na(fu_hrsd_month3 ), fu_hrsd_month3 ,
remapHRSD_QIDS(fu_qids_month3))
			fu_hrsdImputed_month6	<- ifelse(!is.na(fu_hrsd_month6 ), fu_hrsd_month6 ,
remapHRSD_QIDS(fu_qids_month6))
			fu_hrsdImputed_month9	<- ifelse(!is.na(fu_hrsd_month9 ), fu_hrsd_month9 ,
remapHRSD_QIDS(fu_qids_month9))
			fu_hrsdImputed_month12		<- ifelse(!is.na(fu_hrsd_month12 ),
fu_hrsd_month12 , remapHRSD_QIDS(fu_qids_month12))
```

```
# Followup Relapse is defined as any imputed HRSD as >= 14

fu_relapse <- ifelse(  (fu_hrsdImputed_month3 >= 14 & 
!is.na(fu_hrsdImputed_month3)) |

(fu_hrsdImputed_month6 >= 14 & 
!is.na(fu_hrsdImputed_month6)) |

(fu_hrsdImputed_month9 >= 14 & 
!is.na(fu_hrsdImputed_month9)) |

(fu_hrsdImputed_month12 >= 14 & 
!is.na(fu_hrsdImputed_month12)) , 1, 0)


# Followup Sustained remission FULL is defined as all 4 imputed HRSD <= 7

fu_susRemission_full <- ifelse( (fu_hrsdImputed_month3 <= 7 & 
fu_hrsdImputed_month6 <= 7  & fu_hrsdImputed_month9 <= 7 & fu_hrsdImputed_month12 <= 7) &

(!is.na(fu_hrsdImputed_month3) & 
!is.na(fu_hrsdImputed_month6) & !is.na(fu_hrsdImputed_month9) & !is.na(fu_hrsdImputed_month12)), 
1, 0)


# Followup Sustained remission FULL is defined as all 4 imputed HRSD <= 7 
OR MISSING (i.e., no HRSD above > 7)

fu_susRemission_weak  <- ifelse( (fu_hrsdImputed_month3 <= 7  | 
is.na(fu_hrsdImputed_month3)) &

(fu_hrsdImputed_month6 <= 7  | 
is.na(fu_hrsdImputed_month6)) &

(fu_hrsdImputed_month9 <= 7  | 
is.na(fu_hrsdImputed_month9)) &

(fu_hrsdImputed_month12 <= 7  | 
is.na(fu_hrsdImputed_month12)) &

(!is.na(fu_hrsdImputed_month3) | 
!is.na(fu_hrsdImputed_month6) | !is.na(fu_hrsdImputed_month9) | !is.na(fu_hrsdImputed_month12)), 
1, 0)


#Check if they entered remittedi in the maxlevelIVRA before remission


fu_remissionbeforeFU          <- NA

fu_remissionbeforeFU_imputed  <- NA


switch(maxlevel_ivra,

"Level 1" = {fu_remissionbeforeFU            <- 
level1_remitted},

"Level 2" = {fu_remissionbeforeFU            <- 
level2_remitted},

"Level 2A" = {fu_remissionbeforeFU           <- 
level2A_remitted},
```

```
                                    "Level 3" = {fu_remissionbeforeFU            <-
level3_remitted},
                                    "Level 4" = {fu_remissionbeforeFU            <-
level4_remitted}
                              )
                      switch(maxlevel_ivra,
                                    "Level 1" = {fu_remissionbeforeFU_imputed           <-
level1_remitted_imputed},
                                    "Level 2" = {fu_remissionbeforeFU_imputed           <-
level2_remitted_imputed},
                                    "Level 2A" = {fu_remissionbeforeFU_imputed          <-
level2A_remitted_imputed},
                                    "Level 3" = {fu_remissionbeforeFU_imputed           <-
level3_remitted_imputed},
                                    "Level 4" = {fu_remissionbeforeFU_imputed           <-
level4_remitted_imputed}
                              )




              }else if (fu_ivra_entered == 0){

                      fu_hrsd_month3 <- NA
                      fu_hrsd_month6 <- NA
                      fu_hrsd_month9 <- NA
                      fu_hrsd_month12 <- NA
                      fu_qids_month3 <- NA
                      fu_qids_month6 <- NA
                      fu_qids_month9 <- NA
                      fu_qids_month12 <- NA
                      fu_qids_month3_date <- NA
                      fu_qids_month6_date <- NA
                      fu_qids_month9_date <- NA
                      fu_qids_month12_date <- NA

                      fu_hrsdImputed_month3 <- NA
                      fu_hrsdImputed_month6 <- NA
                      fu_hrsdImputed_month9 <- NA
                      fu_hrsdImputed_month12 <- NA
```

```
        fu_relapse  <- NA
        fu_susRemission_full <- NA
        fu_susRemission_weak <- NA
        fu_remissionbeforeFU          <- NA
        fu_remissionbeforeFU_imputed  <- NA

    }

    # Calculate if someone has no followup observations

    # Saving followup outputs

    stardOutputs[stardOutputs$src_subject_id == currentID, "fu_hasHRSDData"] <-
fu_hasHRSDData

    stardOutputs[stardOutputs$src_subject_id == currentID, "fu_hrsd_month3"] <-
fu_hrsd_month3
    stardOutputs[stardOutputs$src_subject_id == currentID, "fu_hrsd_month6"] <-
fu_hrsd_month6
    stardOutputs[stardOutputs$src_subject_id == currentID, "fu_hrsd_month9"] <-
fu_hrsd_month9
    stardOutputs[stardOutputs$src_subject_id == currentID, "fu_hrsd_month12"] <-
fu_hrsd_month12

    stardOutputs[stardOutputs$src_subject_id == currentID, "fu_qids_month3"] <-
fu_qids_month3
    stardOutputs[stardOutputs$src_subject_id == currentID, "fu_qids_month6"] <-
fu_qids_month6
    stardOutputs[stardOutputs$src_subject_id == currentID, "fu_qids_month9"] <-
fu_qids_month9
    stardOutputs[stardOutputs$src_subject_id == currentID, "fu_qids_month12"] <-
fu_qids_month12
    stardOutputs[stardOutputs$src_subject_id == currentID, "fu_qids_month3_date"] <-
fu_qids_month3_date
    stardOutputs[stardOutputs$src_subject_id == currentID, "fu_qids_month6_date"] <-
fu_qids_month6_date
    stardOutputs[stardOutputs$src_subject_id == currentID, "fu_qids_month9_date"] <-
fu_qids_month9_date
    stardOutputs[stardOutputs$src_subject_id == currentID, "fu_qids_month12_date"] <-
fu_qids_month12_date
```

```
                stardOutputs[stardOutputs$src_subject_id == currentID, "fu_hrsdImputed_month3"] <-
fu_hrsdImputed_month3

                stardOutputs[stardOutputs$src_subject_id == currentID, "fu_hrsdImputed_month6"] <-
fu_hrsdImputed_month6

                stardOutputs[stardOutputs$src_subject_id == currentID, "fu_hrsdImputed_month9"] <-
fu_hrsdImputed_month9

                stardOutputs[stardOutputs$src_subject_id == currentID, "fu_hrsdImputed_month12"]
<- fu_hrsdImputed_month12


                stardOutputs[stardOutputs$src_subject_id == currentID, "fu_remissionbeforeFU"]
                <- fu_remissionbeforeFU

                stardOutputs[stardOutputs$src_subject_id == currentID,
"fu_remissionbeforeFU_imputed"]       <- fu_remissionbeforeFU_imputed



                stardOutputs[stardOutputs$src_subject_id == currentID, "fu_relapse"]
        <- fu_relapse

                stardOutputs[stardOutputs$src_subject_id == currentID, "fu_susRemission_full"]
        <- fu_susRemission_full

                stardOutputs[stardOutputs$src_subject_id == currentID, "fu_susRemission_weak"]
        <- fu_susRemission_weak



        print(i)       # Keep track of progress


}



############################################################################################
######################                Cleaning up variables         ######################
############################################################################################



##### Which participants entered each level  ####
```

```
level1_entered3110 <- stardOutputs[stardOutputs$meetInclusion == 1 &
stardOutputs$level1_ivra_entered == 1, "src_subject_id"]

level2_entered1134 <- stardOutputs[stardOutputs$meetInclusion == 1 &
(is.na(stardOutputs$level1_remitted ) | stardOutputs$level1_remitted != 1) &
stardOutputs$level2_ivra_entered == 1, "src_subject_id"]

level2A_entered28 <- stardOutputs[stardOutputs$meetInclusion == 1 &
(is.na(stardOutputs$level1_remitted ) | stardOutputs$level1_remitted != 1) &
(is.na(stardOutputs$level2_remitted ) | stardOutputs$level2_remitted != 1) &
stardOutputs$level2A_ivra_entered == 1, "src_subject_id"]

level3_entered313 <- stardOutputs[stardOutputs$meetInclusion == 1 &
(is.na(stardOutputs$level1_remitted ) | stardOutputs$level1_remitted != 1) &
(is.na(stardOutputs$level2_remitted ) | stardOutputs$level2_remitted != 1) &
(is.na(stardOutputs$level2A_remitted ) | stardOutputs$level2A_remitted != 1)  &
stardOutputs$level3_ivra_entered == 1, "src_subject_id"]

level4_entered94 <- stardOutputs[stardOutputs$meetInclusion == 1 &
(is.na(stardOutputs$level1_remitted ) | stardOutputs$level1_remitted != 1) &
(is.na(stardOutputs$level2_remitted ) | stardOutputs$level2_remitted != 1) &
(is.na(stardOutputs$level2A_remitted ) | stardOutputs$level2A_remitted != 1)  &
(is.na(stardOutputs$level3_remitted ) | stardOutputs$level3_remitted != 1)  &
stardOutputs$level4_ivra_entered == 1, "src_subject_id"]




############ Create function to calculate mean, 95% CI's and SD's   ###############


summarizeOutputs       <- function(x){

        meanX <- mean(x, na.rm = T)

        nX      <- sum(!is.na(x))

        sdX     <- sd(x, na.rm = T)

        seX     <- sdX/sqrt(nX)

        upper95X       <- meanX + 1.96*seX

        lower95X       <- meanX  - 1.96*seX


        summaryOutput <- c(mean = meanX,

                                              sd = sdX,

                                              upper95 = upper95X,

                                              lower95 = lower95X)

        return(summaryOutput)


}
```

```
################################          Calculations for followup     ##################


######   Create variable for people who have no IVRA followup assessments   ##########


# This is how many people entered followup but had NO measurements

stardOutputs$fu_noObservations <- with(stardOutputs, ifelse(is.na(fu_hrsd_month3) &
is.na(fu_hrsd_month6) & is.na(fu_hrsd_month9) & is.na(fu_hrsd_month12)

                                                    & is.na(fu_qids_month3) &
is.na(fu_qids_month6) & is.na(fu_qids_month9) & is.na(fu_qids_month12), 1,0))




# This is how many people entered followup and had at least ONE measurement

stardOutputs$fu_hasAtLeast1Observations <- with(stardOutputs, ifelse(!is.na(fu_hrsd_month3) |
!is.na(fu_hrsd_month6) | !is.na(fu_hrsd_month9) | !is.na(fu_hrsd_month12)

                                                    | !is.na(fu_qids_month3) |
!is.na(fu_qids_month6) | !is.na(fu_qids_month9) | !is.na(fu_qids_month12), 1,0))





table(stardOutputs[stardOutputs$meetInclusion == 1 & stardOutputs$fu_ivra_entered ==
1,]$fu_noObservations )

table(stardOutputs[stardOutputs$meetInclusion == 1 & stardOutputs$fu_ivra_entered ==
1,]$fu_hasAtLeast1Observations )




##### Determining who entered followup before remission            ####


# People who entered followup with remission, and then later met relpase



stardOutputs$fu_remit_then_relapse                   <-
ifelse(stardOutputs$fu_remissionbeforeFU== 1 &      stardOutputs$fu_relapse ==1, 1,0)

stardOutputs$fu_NOremit_then_relapse                 <-
ifelse(stardOutputs$fu_remissionbeforeFU== 0 &      stardOutputs$fu_relapse ==1, 1,0)

stardOutputs$fu_remitImputed_then_relapse            <-
ifelse(stardOutputs$fu_remissionbeforeFU_imputed == 1 &     stardOutputs$fu_relapse ==1, 1,0)

stardOutputs$fu_NOremitImputed_then_relapse          <-
ifelse(stardOutputs$fu_remissionbeforeFU_imputed == 0 &     stardOutputs$fu_relapse ==1, 1,0)




stardOutputs$fu_remit_then_susRemission_full                      <-
ifelse(stardOutputs$fu_remissionbeforeFU == 1 &     stardOutputs$fu_susRemission_full ==1, 1,0)
```

```
stardOutputs$fu_NOremit_then_susRemission_full              <-
ifelse(stardOutputs$fu_remissionbeforeFU == 0 &      stardOutputs$fu_susRemission_full ==1, 1,0)

stardOutputs$fu_remitImputed_then_susRemission_full          <-
ifelse(stardOutputs$fu_remissionbeforeFU_imputed == 1 &     stardOutputs$fu_susRemission_full
==1, 1,0)

stardOutputs$fu_NOremitImputed_then_susRemission_full        <-
ifelse(stardOutputs$fu_remissionbeforeFU_imputed == 0 &     stardOutputs$fu_susRemission_full
==1, 1,0)




stardOutputs$fu_remit_then_susRemission_weak                        <-
ifelse(stardOutputs$fu_remissionbeforeFU == 1 &      stardOutputs$fu_susRemission_weak ==1, 1,0)

stardOutputs$fu_NOremit_then_susRemission_weak              <-
ifelse(stardOutputs$fu_remissionbeforeFU == 0 &      stardOutputs$fu_susRemission_weak ==1, 1,0)

stardOutputs$fu_remitImputed_then_susRemission_weak          <-
ifelse(stardOutputs$fu_remissionbeforeFU_imputed == 1 &     stardOutputs$fu_susRemission_weak
==1, 1,0)

stardOutputs$fu_NOremitImputed_then_susRemission_weak        <-
ifelse(stardOutputs$fu_remissionbeforeFU_imputed == 0 &     stardOutputs$fu_susRemission_weak
==1, 1,0)




##########################                   Statistical Tests      Comparisons   #############


########       ANOVA To compare mean HRSD Change among Level 2 treatments



level2SwitchPatients   <- stardOutputs[stardOutputs$src_subject_id %in% level2_entered1134 &
stardOutputs$level2_tx %in% c("SER", "VEN", "BUP"),]


level2AugmentationData <- stardOutputs[stardOutputs$src_subject_id %in% level2_entered1134 &
stardOutputs$level2_tx %in% c("CIT+BUP", "CIT+BUS", "CIT+CT"),]


summary(aov(lm(level2_hrsdchange ~ level2_tx, data = level2SwitchPatients)))

summary(aov(lm(level2_hrsdchange_imputed ~ level2_tx, data = level2SwitchPatients)))


summary(aov(lm(level2_hrsdchange ~ level2_tx, data = level2AugmentationData)))

summary(aov(lm(level2_hrsdchange_imputed ~ level2_tx, data = level2AugmentationData)))
```

```
####################################################################################################
####
######################                OUTPUTS                               ######################
####################################################################################################
####

## The max level accoring to each person on IVRA
table(stardOutputs$maxlevel_ivra)
table(stardOutputs[stardOutputs$meetInclusion == 1,]$maxlevel_ivra)

###            Calculating how many meet HRSD criteria at DaysBaseline <=5                ####

#How many HRSD ≤ 7
table(stardOutputs$level1_first_hrsd_sum     <= 7 , useNA = "always")

#How many 7 < HRSD < 14
table(stardOutputs$level1_first_hrsd_sum<  14 & stardOutputs$level1_first_hrsd_sum> 7 , useNA =
"always")

## How many have missing hdtot_sum_NArmT at enrollment
table(is.na(stardOutputs$level1_first_hrsd_sum), useNA = "always")

#####   How many meet inclusion       ###

table(stardOutputs$meetInclusion, useNA = "always")

## How many people entered which level
table(stardOutputs[stardOutputs$meetInclusion == 1,]$level1_ivra_entered, useNA = "always")
table(stardOutputs[stardOutputs$meetInclusion == 1,]$level2_ivra_entered, useNA = "always")
table(stardOutputs[stardOutputs$meetInclusion == 1,]$level2A_ivra_entered, useNA = "always")
table(stardOutputs[stardOutputs$meetInclusion == 1,]$level3_ivra_entered, useNA = "always")
```

```
table(stardOutputs[stardOutputs$meetInclusion == 1,]$level4_ivra_entered, useNA = "always")

## How many people entered each level according to IVRA, and met inclusion criteria

nrow(stardOutputs[stardOutputs$meetInclusion == 1 & stardOutputs$level1_ivra_entered == 1,])

nrow(stardOutputs[stardOutputs$meetInclusion == 1 & stardOutputs$level2_ivra_entered == 1,])

nrow(stardOutputs[stardOutputs$meetInclusion == 1 & stardOutputs$level2A_ivra_entered == 1,])

nrow(stardOutputs[stardOutputs$meetInclusion == 1 & stardOutputs$level3_ivra_entered == 1,])

nrow(stardOutputs[stardOutputs$meetInclusion == 1 & stardOutputs$level4_ivra_entered == 1,])

## How many people entered each level according to IVRA, and did not meet remission for previous
level , and met inclusion criteria

nrow(stardOutputs[stardOutputs$meetInclusion == 1 & stardOutputs$level1_ivra_entered == 1,])

nrow(stardOutputs[stardOutputs$meetInclusion == 1 & (is.na(stardOutputs$level1_remitted ) |
stardOutputs$level1_remitted != 1) & stardOutputs$level2_ivra_entered == 1,])

nrow(stardOutputs[stardOutputs$meetInclusion == 1 & (is.na(stardOutputs$level1_remitted ) |
stardOutputs$level1_remitted != 1) & (is.na(stardOutputs$level2_remitted ) |
stardOutputs$level2_remitted != 1) & stardOutputs$level2A_ivra_entered == 1,])

nrow(stardOutputs[stardOutputs$meetInclusion == 1 & (is.na(stardOutputs$level1_remitted ) |
stardOutputs$level1_remitted != 1) & (is.na(stardOutputs$level2_remitted ) |
stardOutputs$level2_remitted != 1) & (is.na(stardOutputs$level2A_remitted ) |
stardOutputs$level2A_remitted != 1)  & stardOutputs$level3_ivra_entered == 1,])

nrow(stardOutputs[stardOutputs$meetInclusion == 1 & (is.na(stardOutputs$level1_remitted ) |
stardOutputs$level1_remitted != 1) & (is.na(stardOutputs$level2_remitted ) |
stardOutputs$level2_remitted != 1) & (is.na(stardOutputs$level2A_remitted ) |
stardOutputs$level2A_remitted != 1)  & (is.na(stardOutputs$level3_remitted ) |
stardOutputs$level3_remitted != 1)  & stardOutputs$level4_ivra_entered == 1,])

## These are how many people entered each level by treatment

with(stardOutputs[stardOutputs$meetInclusion == 1,] , table(level1_tx, useNA = "always"))

with(stardOutputs[stardOutputs$meetInclusion == 1 & (is.na(stardOutputs$level1_remitted ) |
stardOutputs$level1_remitted != 1),] , table(level2_tx, useNA = "always"))

with(stardOutputs[stardOutputs$meetInclusion == 1 & (is.na(stardOutputs$level1_remitted ) |
stardOutputs$level1_remitted != 1) & (is.na(stardOutputs$level2_remitted ) |
stardOutputs$level2_remitted != 1) & stardOutputs$level2A_ivra_entered == 1,], table(level2A_tx,
useNA = "always"))

with(stardOutputs[stardOutputs$meetInclusion == 1 & (is.na(stardOutputs$level1_remitted ) |
stardOutputs$level1_remitted != 1) & (is.na(stardOutputs$level2_remitted ) |
stardOutputs$level2_remitted != 1) & (is.na(stardOutputs$level2A_remitted ) |
stardOutputs$level2A_remitted != 1)  & stardOutputs$level3_ivra_entered == 1,], table(level3_tx,
useNA = "always"))
```

```
with(stardOutputs[stardOutputs$meetInclusion == 1 & (is.na(stardOutputs$level1_remitted ) |
stardOutputs$level1_remitted != 1) & (is.na(stardOutputs$level2_remitted ) |
stardOutputs$level2_remitted != 1) & (is.na(stardOutputs$level2A_remitted ) |
stardOutputs$level2A_remitted != 1)  & (is.na(stardOutputs$level3_remitted ) |
stardOutputs$level3_remitted != 1)  & stardOutputs$level4_ivra_entered == 1,], table(level4_tx,
useNA = "always"))


## Now find remissions for each level by treatments ##


with(stardOutputs[stardOutputs$meetInclusion == 1 & stardOutputs$level1_ivra_entered == 1,],
table(level1_remitted, level1_tx, useNA = "always"))

with(stardOutputs[stardOutputs$meetInclusion == 1 & (is.na(stardOutputs$level1_remitted ) |
stardOutputs$level1_remitted != 1) & stardOutputs$level2_ivra_entered == 1,],
table(level2_remitted, level2_tx, useNA = "always"))

with(stardOutputs[stardOutputs$meetInclusion == 1 & (is.na(stardOutputs$level1_remitted ) |
stardOutputs$level1_remitted != 1) & (is.na(stardOutputs$level2_remitted ) |
stardOutputs$level2_remitted != 1) & stardOutputs$level2A_ivra_entered == 1,],
table(level2A_remitted, level2A_tx, useNA = "always"))

with(stardOutputs[stardOutputs$meetInclusion == 1  & (is.na(stardOutputs$level1_remitted ) |
stardOutputs$level1_remitted != 1) & (is.na(stardOutputs$level2_remitted ) |
stardOutputs$level2_remitted != 1) & (is.na(stardOutputs$level2A_remitted ) |
stardOutputs$level2A_remitted != 1)  & stardOutputs$level3_ivra_entered == 1,],
table(level3_remitted, level3_tx, useNA = "always"))

with(stardOutputs[stardOutputs$meetInclusion == 1 & (is.na(stardOutputs$level1_remitted ) |
stardOutputs$level1_remitted != 1) & (is.na(stardOutputs$level2_remitted ) |
stardOutputs$level2_remitted != 1) & (is.na(stardOutputs$level2A_remitted ) |
stardOutputs$level2A_remitted != 1)  & (is.na(stardOutputs$level3_remitted ) |
stardOutputs$level3_remitted != 1)  & stardOutputs$level4_ivra_entered == 1,],
table(level4_remitted, level4_tx, useNA = "always"))


## Remission rates with imputed QIDS


with(stardOutputs[stardOutputs$meetInclusion == 1 & stardOutputs$level1_ivra_entered == 1,],
table(level1_remitted_imputed , level1_tx, useNA = "always"))

with(stardOutputs[stardOutputs$meetInclusion == 1 & (is.na(stardOutputs$level1_remitted ) |
stardOutputs$level1_remitted != 1) & stardOutputs$level2_ivra_entered == 1,],
table(level2_remitted_imputed, level2_tx, useNA = "always"))

with(stardOutputs[stardOutputs$meetInclusion == 1 & (is.na(stardOutputs$level1_remitted ) |
stardOutputs$level1_remitted != 1) & (is.na(stardOutputs$level2_remitted ) |
stardOutputs$level2_remitted != 1) & stardOutputs$level2A_ivra_entered == 1,],
table(level2A_remitted_imputed, level2A_tx, useNA = "always"))

with(stardOutputs[stardOutputs$meetInclusion == 1  & (is.na(stardOutputs$level1_remitted ) |
stardOutputs$level1_remitted != 1) & (is.na(stardOutputs$level2_remitted ) |
stardOutputs$level2_remitted != 1) & (is.na(stardOutputs$level2A_remitted ) |
stardOutputs$level2A_remitted != 1)  & stardOutputs$level3_ivra_entered == 1,],
table(level3_remitted_imputed, level3_tx, useNA = "always"))

with(stardOutputs[stardOutputs$meetInclusion == 1 & (is.na(stardOutputs$level1_remitted ) |
stardOutputs$level1_remitted != 1) & (is.na(stardOutputs$level2_remitted ) |
stardOutputs$level2_remitted != 1) & (is.na(stardOutputs$level2A_remitted ) |
```

```
stardOutputs$level2A_remitted != 1)  & (is.na(stardOutputs$level3_remitted ) |
stardOutputs$level3_remitted != 1)  & stardOutputs$level4_ivra_entered == 1,],
table(level4_remitted_imputed, level4_tx, useNA = "always"))
```

```
## Now find responses for each level by treatments ##
```

```
with(stardOutputs[stardOutputs$meetInclusion == 1 & stardOutputs$level1_ivra_entered == 1,],
table(level1_responded, level1_tx, useNA = "always"))

with(stardOutputs[stardOutputs$meetInclusion == 1 & (is.na(stardOutputs$level1_remitted ) |
stardOutputs$level1_remitted != 1) & stardOutputs$level2_ivra_entered == 1,],
table(level2_responded, level2_tx, useNA = "always"))

with(stardOutputs[stardOutputs$meetInclusion == 1 & (is.na(stardOutputs$level1_remitted ) |
stardOutputs$level1_remitted != 1) & (is.na(stardOutputs$level2_remitted ) |
stardOutputs$level2_remitted != 1) & stardOutputs$level2A_ivra_entered == 1,],
table(level2A_responded, level2A_tx, useNA = "always"))

with(stardOutputs[stardOutputs$meetInclusion == 1  & (is.na(stardOutputs$level1_remitted ) |
stardOutputs$level1_remitted != 1) & (is.na(stardOutputs$level2_remitted ) |
stardOutputs$level2_remitted != 1) & (is.na(stardOutputs$level2A_remitted ) |
stardOutputs$level2A_remitted != 1)  & stardOutputs$level3_ivra_entered == 1,],
table(level3_responded, level3_tx, useNA = "always"))

with(stardOutputs[stardOutputs$meetInclusion == 1 & (is.na(stardOutputs$level1_remitted ) |
stardOutputs$level1_remitted != 1) & (is.na(stardOutputs$level2_remitted ) |
stardOutputs$level2_remitted != 1) & (is.na(stardOutputs$level2A_remitted ) |
stardOutputs$level2A_remitted != 1)  & (is.na(stardOutputs$level3_remitted ) |
stardOutputs$level3_remitted != 1)  & stardOutputs$level4_ivra_entered == 1,],
table(level4_responded, level4_tx, useNA = "always"))
```

```
## Now find imputed responses for each level by treatments ##
```

```
with(stardOutputs[stardOutputs$meetInclusion == 1 & stardOutputs$level1_ivra_entered == 1,],
table(level1_responded_imputed, level1_tx, useNA = "always"))

with(stardOutputs[stardOutputs$meetInclusion == 1 & (is.na(stardOutputs$level1_remitted ) |
stardOutputs$level1_remitted != 1) & stardOutputs$level2_ivra_entered == 1,],
table(level2_responded_imputed, level2_tx, useNA = "always"))

with(stardOutputs[stardOutputs$meetInclusion == 1 & (is.na(stardOutputs$level1_remitted ) |
stardOutputs$level1_remitted != 1) & (is.na(stardOutputs$level2_remitted ) |
stardOutputs$level2_remitted != 1) & stardOutputs$level2A_ivra_entered == 1,],
table(level2A_responded_imputed, level2A_tx, useNA = "always"))

with(stardOutputs[stardOutputs$meetInclusion == 1  & (is.na(stardOutputs$level1_remitted ) |
stardOutputs$level1_remitted != 1) & (is.na(stardOutputs$level2_remitted ) |
stardOutputs$level2_remitted != 1) & (is.na(stardOutputs$level2A_remitted ) |
stardOutputs$level2A_remitted != 1)  & stardOutputs$level3_ivra_entered == 1,],
table(level3_responded_imputed, level3_tx, useNA = "always"))

with(stardOutputs[stardOutputs$meetInclusion == 1 & (is.na(stardOutputs$level1_remitted ) |
stardOutputs$level1_remitted != 1) & (is.na(stardOutputs$level2_remitted ) |
stardOutputs$level2_remitted != 1) & (is.na(stardOutputs$level2A_remitted ) |
stardOutputs$level2A_remitted != 1)  & (is.na(stardOutputs$level3_remitted ) |
stardOutputs$level3_remitted != 1)  & stardOutputs$level4_ivra_entered == 1,],
table(level4_responded_imputed, level4_tx, useNA = "always"))
```

```
## Now find mean HRSD change score  ##



with(stardOutputs[stardOutputs$meetInclusion == 1 & stardOutputs$level1_ivra_entered == 1,],
aggregate(level1_hrsdchange, list(level1_tx), mean, na.rm = T))

with(stardOutputs[stardOutputs$meetInclusion == 1 & (is.na(stardOutputs$level1_remitted ) |
stardOutputs$level1_remitted != 1) & stardOutputs$level2_ivra_entered == 1,],
aggregate(level2_hrsdchange, list(level2_tx), mean, na.rm = T))

with(stardOutputs[stardOutputs$meetInclusion == 1 & (is.na(stardOutputs$level1_remitted ) |
stardOutputs$level1_remitted != 1) & (is.na(stardOutputs$level2_remitted ) |
stardOutputs$level2_remitted != 1) & stardOutputs$level2A_ivra_entered == 1,],
aggregate(level2A_hrsdchange, list(level2A_tx), mean, na.rm = T))

with(stardOutputs[stardOutputs$meetInclusion == 1  & (is.na(stardOutputs$level1_remitted ) |
stardOutputs$level1_remitted != 1) & (is.na(stardOutputs$level2_remitted ) |
stardOutputs$level2_remitted != 1) & (is.na(stardOutputs$level2A_remitted ) |
stardOutputs$level2A_remitted != 1)  & stardOutputs$level3_ivra_entered == 1,],
aggregate(level3_hrsdchange, list(level3_tx), mean, na.rm = T))

with(stardOutputs[stardOutputs$meetInclusion == 1 & (is.na(stardOutputs$level1_remitted ) |
stardOutputs$level1_remitted != 1) & (is.na(stardOutputs$level2_remitted ) |
stardOutputs$level2_remitted != 1) & (is.na(stardOutputs$level2A_remitted ) |
stardOutputs$level2A_remitted != 1)  & (is.na(stardOutputs$level3_remitted ) |
stardOutputs$level3_remitted != 1)  & stardOutputs$level4_ivra_entered == 1,],
aggregate(level4_hrsdchange, list(level4_tx), mean, na.rm = T))



## Now find mean HRSD change imputed score  ##



with(stardOutputs[stardOutputs$meetInclusion == 1 & stardOutputs$level1_ivra_entered == 1,],
aggregate(level1_hrsdchange_imputed, list(level1_tx), summarizeOutputs))

with(stardOutputs[stardOutputs$meetInclusion == 1 & (is.na(stardOutputs$level1_remitted ) |
stardOutputs$level1_remitted != 1) & stardOutputs$level2_ivra_entered == 1,],
aggregate(level2_hrsdchange_imputed, list(level2_tx),  summarizeOutputs))

with(stardOutputs[stardOutputs$meetInclusion == 1 & (is.na(stardOutputs$level1_remitted ) |
stardOutputs$level1_remitted != 1) & (is.na(stardOutputs$level2_remitted ) |
stardOutputs$level2_remitted != 1) & stardOutputs$level2A_ivra_entered == 1,],
aggregate(level2A_hrsdchange_imputed, list(level2A_tx),  summarizeOutputs))

with(stardOutputs[stardOutputs$meetInclusion == 1  & (is.na(stardOutputs$level1_remitted ) |
stardOutputs$level1_remitted != 1) & (is.na(stardOutputs$level2_remitted ) |
stardOutputs$level2_remitted != 1) & (is.na(stardOutputs$level2A_remitted ) |
stardOutputs$level2A_remitted != 1)  & stardOutputs$level3_ivra_entered == 1,],
aggregate(level3_hrsdchange_imputed, list(level3_tx),  summarizeOutputs))

with(stardOutputs[stardOutputs$meetInclusion == 1 & (is.na(stardOutputs$level1_remitted ) |
stardOutputs$level1_remitted != 1) & (is.na(stardOutputs$level2_remitted ) |
stardOutputs$level2_remitted != 1) & (is.na(stardOutputs$level2A_remitted ) |
stardOutputs$level2A_remitted != 1)  & (is.na(stardOutputs$level3_remitted ) |
stardOutputs$level3_remitted != 1)  & stardOutputs$level4_ivra_entered == 1,],
aggregate(level4_hrsdchange_imputed, list(level4_tx),  summarizeOutputs))
```

```
### Find mean HRSD    ##


with(stardOutputs[stardOutputs$meetInclusion == 1 & stardOutputs$level1_ivra_entered == 1,],
aggregate(level1_last_hrsd_sum, list(level1_tx), mean, na.rm = T))

with(stardOutputs[stardOutputs$meetInclusion == 1 & (is.na(stardOutputs$level1_remitted ) |
stardOutputs$level1_remitted != 1) & stardOutputs$level2_ivra_entered == 1,],
aggregate(level1_last_hrsd_sum, list(level2_tx), mean, na.rm = T))

with(stardOutputs[stardOutputs$meetInclusion == 1 & (is.na(stardOutputs$level1_remitted ) |
stardOutputs$level1_remitted != 1) & stardOutputs$level2_ivra_entered == 1,],
aggregate(level1_last_hrsd_sum, list(level2_tx), sd, na.rm = T))


with(stardOutputs[stardOutputs$meetInclusion == 1 & (is.na(stardOutputs$level1_remitted ) |
stardOutputs$level1_remitted != 1) & stardOutputs$level2_ivra_entered == 1

               & stardOutputs$level2_tx %in% c("BUP", "SER", "VEN"),],
aggregate(level1_last_hrsd_sum, list(level2_tx), mean, na.rm = T))


with(stardOutputs[stardOutputs$meetInclusion == 1 & (is.na(stardOutputs$level1_remitted ) |
stardOutputs$level1_remitted != 1) & stardOutputs$level2_ivra_entered == 1

               & stardOutputs$level2_tx %in% c("BUP", "SER", "VEN"),], mean(level1_last_hrsd_sum,
na.rm = T))

with(stardOutputs[stardOutputs$meetInclusion == 1 & (is.na(stardOutputs$level1_remitted ) |
stardOutputs$level1_remitted != 1) & stardOutputs$level2_ivra_entered == 1

               & stardOutputs$level2_tx %in% c("BUP", "SER", "VEN"),], sd(level1_last_hrsd_sum,
na.rm = T))


with(stardOutputs[stardOutputs$meetInclusion == 1 & (is.na(stardOutputs$level1_remitted ) |
stardOutputs$level1_remitted != 1) & stardOutputs$level2_ivra_entered == 1

               & stardOutputs$level2_tx %in% c("CIT+BUP", "CIT+BUS"),],
mean(level1_last_hrsd_sum, na.rm = T))

with(stardOutputs[stardOutputs$meetInclusion == 1 & (is.na(stardOutputs$level1_remitted ) |
stardOutputs$level1_remitted != 1) & stardOutputs$level2_ivra_entered == 1

               & stardOutputs$level2_tx %in% c("CIT+BUP", "CIT+BUS"),], sd(level1_last_hrsd_sum,
na.rm = T))


################################################################################################
#################################

############################                         FOLLOWUP CALCULATIONS
       ########################################

################################################################################################
#################################


## Who entered according to IVRA each level  ##
```

```
with(stardOutputs[stardOutputs$meetInclusion == 1,], table(level1_ivra_entered, useNA =
"always"))

with(stardOutputs[stardOutputs$meetInclusion == 1 & (stardOutputs$level1_remitted !=  1  |
is.na(stardOutputs$level1_remitted)),], table(level2_ivra_entered, useNA = "always"))

with(stardOutputs[stardOutputs$meetInclusion == 1 & (stardOutputs$level1_remitted !=  1  |
is.na(stardOutputs$level1_remitted)) & (stardOutputs$level2_remitted !=  1  |
is.na(stardOutputs$level2_remitted)),], table(level2A_ivra_entered, useNA = "always"))

with(stardOutputs[stardOutputs$meetInclusion == 1 & (is.na(stardOutputs$level1_remitted ) |
stardOutputs$level1_remitted != 1) & (is.na(stardOutputs$level2_remitted ) |
stardOutputs$level2_remitted != 1) & (is.na(stardOutputs$level2A_remitted ) |
stardOutputs$level2A_remitted != 1),], table(level3_ivra_entered , useNA = "always"))

with(stardOutputs[stardOutputs$meetInclusion == 1 & (is.na(stardOutputs$level1_remitted ) |
stardOutputs$level1_remitted != 1) & (is.na(stardOutputs$level2_remitted ) |
stardOutputs$level2_remitted != 1) & (is.na(stardOutputs$level2A_remitted ) |
stardOutputs$level2A_remitted != 1)  & (is.na(stardOutputs$level3_remitted ) |
stardOutputs$level3_remitted != 1),], table(level4_ivra_entered , useNA = "always"))



## Max level IVRA for each    ##

with(stardOutputs, table(maxlevel_ivra, useNA = "always"))

with(stardOutputs[stardOutputs$meetInclusion == 1,], table(maxlevel_ivra, useNA = "always"))



##################



## How many entered followup in each level according to IVRA - This preserves the correct
denominator and uses whether they have followup IVRA

with(stardOutputs[stardOutputs$meetInclusion == 1 & !(stardOutputs$src_subject_id %in%
c(level2_entered1134 , level2A_entered28, level3_entered313, level4_entered94)),],
table(fu_ivra_entered, useNA = "always"))

with(stardOutputs[stardOutputs$src_subject_id %in% level2_entered1134 &
!(stardOutputs$src_subject_id %in% c(level2A_entered28, level3_entered313, level4_entered94)),],
table(fu_ivra_entered, useNA = "always"))

with(stardOutputs[stardOutputs$src_subject_id %in% level2A_entered28 &
!(stardOutputs$src_subject_id %in% c(level3_entered313, level4_entered94)),],
table(fu_ivra_entered, useNA = "always"))

with(stardOutputs[stardOutputs$src_subject_id %in% level3_entered313 &
!(stardOutputs$src_subject_id %in% c(level4_entered94)),], table(fu_ivra_entered, useNA =
"always"))

with(stardOutputs[stardOutputs$src_subject_id %in% level4_entered94,], table(fu_ivra_entered,
useNA = "always"))



## How many entered followup in each level according to IVRA - This preserves the correct
denominator and uses whether they have followup IVRA
```

```
with(stardOutputs[stardOutputs$meetInclusion == 1 & !(stardOutputs$src_subject_id %in%
c(level2_entered1134 , level2A_entered28, level3_entered313, level4_entered94)),],
table(fu_ivra_entered, useNA = "always"))

with(stardOutputs[stardOutputs$src_subject_id %in% level2_entered1134 &
!(stardOutputs$src_subject_id %in% c(level2A_entered28, level3_entered313, level4_entered94)),],
table(fu_ivra_entered, useNA = "always"))

with(stardOutputs[stardOutputs$src_subject_id %in% level2A_entered28 &
stardOutputs$level1_remitted != 1  & !(stardOutputs$src_subject_id %in% c(level3_entered313,
level4_entered94)),], table(fu_ivra_entered, useNA = "always"))

with(stardOutputs[stardOutputs$src_subject_id %in% level3_entered313 &
stardOutputs$level1_remitted != 1  & !(stardOutputs$src_subject_id %in% c(level4_entered94)),],
table(fu_ivra_entered, useNA = "always"))

with(stardOutputs[stardOutputs$src_subject_id %in% level4_entered94,], table(fu_ivra_entered,
useNA = "always"))
```

```
## How many have followup HRSD at each level - This preserves the correct deminator and uses
whether they have followup HRSD

with(stardOutputs[stardOutputs$meetInclusion == 1 & !(stardOutputs$src_subject_id %in%
c(level2_entered1134 , level2A_entered28, level3_entered313, level4_entered94)),],
table(fu_hasHRSDData , useNA = "always"))

with(stardOutputs[stardOutputs$src_subject_id %in% level2_entered1134 &
!(stardOutputs$src_subject_id %in% c(level2A_entered28, level3_entered313, level4_entered94)),],
table(fu_hasHRSDData , useNA = "always"))

with(stardOutputs[stardOutputs$src_subject_id %in% level2A_entered28 &
!(stardOutputs$src_subject_id %in% c(level3_entered313, level4_entered94)),],
table(fu_hasHRSDData , useNA = "always"))

with(stardOutputs[stardOutputs$src_subject_id %in% level3_entered313 &
!(stardOutputs$src_subject_id %in% c(level4_entered94)),], table(fu_hasHRSDData , useNA =
"always"))

with(stardOutputs[stardOutputs$src_subject_id %in% level4_entered94,], table(fu_hasHRSDData ,
useNA = "always"))
```

```
## How many hve followup HRSD according to max IVRA -- This may lead to inconsistencies, because
individuals could have been incorrectly moved on via IVRA despite meeting remission on HRSD.

with(stardOutputs[stardOutputs$meetInclusion == 1 & stardOutputs$maxlevel_ivra == "Level 1",],
table(fu_hasHRSDData, useNA = "always"))

with(stardOutputs[stardOutputs$meetInclusion == 1 & stardOutputs$maxlevel_ivra == "Level 2",],
table(fu_hasHRSDData, useNA = "always"))

with(stardOutputs[stardOutputs$meetInclusion == 1 & stardOutputs$maxlevel_ivra == "Level 2A",],
table(fu_hasHRSDData, useNA = "always"))

with(stardOutputs[stardOutputs$meetInclusion == 1 & stardOutputs$maxlevel_ivra == "Level 3",],
table(fu_hasHRSDData, useNA = "always"))

with(stardOutputs[stardOutputs$meetInclusion == 1 & stardOutputs$maxlevel_ivra == "Level 4",],
table(fu_hasHRSDData, useNA = "always"))
```

```
###### Calculating sustained remission       ########




## How many entered followup in each level according to IVRA - This preserves the correct denominator and uses whether they have followup IVRA

with(stardOutputs[stardOutputs$meetInclusion == 1 & !(stardOutputs$src_subject_id %in% c(level2_entered1134 , level2A_entered28, level3_entered313, level4_entered94)),], table(fu_ivra_entered, level1_tx, useNA = "always"))

with(stardOutputs[stardOutputs$meetInclusion == 1 & !(stardOutputs$src_subject_id %in% c(level2_entered1134 , level2A_entered28, level3_entered313, level4_entered94)),], table(fu_relapse, level1_tx, useNA = "always"))

with(stardOutputs[stardOutputs$meetInclusion == 1 & !(stardOutputs$src_subject_id %in% c(level2_entered1134 , level2A_entered28, level3_entered313, level4_entered94)),], table(fu_susRemission_full, level1_tx, useNA = "always"))

with(stardOutputs[stardOutputs$meetInclusion == 1 & !(stardOutputs$src_subject_id %in% c(level2_entered1134 , level2A_entered28, level3_entered313, level4_entered94)),], table(fu_susRemission_weak, level1_tx, useNA = "always"))



with(stardOutputs[stardOutputs$src_subject_id %in% level2_entered1134 & !(stardOutputs$src_subject_id %in% c(level2A_entered28, level3_entered313, level4_entered94)),], table(fu_ivra_entered, level2_tx, useNA = "always"))

with(stardOutputs[stardOutputs$src_subject_id %in% level2_entered1134 & !(stardOutputs$src_subject_id %in% c(level2A_entered28, level3_entered313, level4_entered94)),], table(fu_relapse, level2_tx, useNA = "always"))

with(stardOutput      s[stardOutputs$src_subject_id %in% level2_entered1134 & !(stardOutputs$src_subject_id %in% c(level2A_entered28, level3_entered313, level4_entered94)),], table(fu_susRemission_full, level2_tx, useNA = "always"))

with(stardOutputs[stardOutputs$src_subject_id %in% level2_entered1134 & !(stardOutputs$src_subject_id %in% c(level2A_entered28, level3_entered313, level4_entered94)),], table(fu_susRemission_weak, level2_tx, useNA = "always"))



with(stardOutputs[stardOutputs$src_subject_id %in% level2A_entered28 & !(stardOutputs$src_subject_id %in% c(level3_entered313, 13, level4_entered94)),], table(fu_ivra_entered, level2A_tx, useNA = "always"))

with(stardOutputs[stardOutputs$src_subject_id %in% level2A_entered28 & !(stardOutputs$src_subject_id %in% c(level3_entered313, 13, level4_entered94)),], table(fu_relapse, level2A_tx, useNA = "always"))

with(stardOutputs[stardOutputs$src_subject_id %in% level2A_entered28 & !(stardOutputs$src_subject_id %in% c(level3_entered313, 13, level4_entered94)),], table(fu_susRemission_full, level2A_tx, useNA = "always"))

with(stardOutputs[stardOutputs$src_subject_id %in% level2A_entered28 & !(stardOutputs$src_subject_id %in% c(level3_entered313, 13, level4_entered94)),], table(fu_susRemission_weak, level2A_tx, useNA = "always"))
```

```
with(stardOutputs[stardOutputs$src_subject_id %in% level3_entered313 &
!(stardOutputs$src_subject_id %in% c(level4_entered94)),], table(fu_ivra_entered, level3_tx,
useNA = "always"))

with(stardOutputs[stardOutputs$src_subject_id %in% level3_entered313 &
!(stardOutputs$src_subject_id %in% c(level4_entered94)),], table(fu_relapse, level3_tx, useNA =
"always"))

with(stardOutputs[stardOutputs$src_subject_id %in% level3_entered313 &
!(stardOutputs$src_subject_id %in% c(level4_entered94)),], table(fu_susRemission_full, level3_tx,
useNA = "always"))

with(stardOutputs[stardOutputs$src_subject_id %in% level3_entered313 &
!(stardOutputs$src_subject_id %in% c(level4_entered94)),], table(fu_susRemission_weak, level3_tx,
useNA = "always"))


with(stardOutputs[stardOutputs$src_subject_id %in% level4_entered94,], table(fu_ivra_entered,
level4_tx, useNA = "always"))

with(stardOutputs[stardOutputs$src_subject_id %in% level4_entered94,], table(fu_relapse,
level4_tx, useNA = "always"))

with(stardOutputs[stardOutputs$src_subject_id %in% level4_entered94,],
table(fu_susRemission_full, level4_tx, useNA = "always"))

with(stardOutputs[stardOutputs$src_subject_id %in% level4_entered94,],
table(fu_susRemission_weak, level4_tx, useNA = "always"))


#################################################################################################
########################

########################                    COMBINING INTO A SINGLE OUTPUT
       ################################

#################################################################################################
########################


## Level 1     ##

(outputs_level1_count                       <- with(stardOutputs[stardOutputs$meetInclusion ==
1,] , table(level1_tx, useNA = "always")))

(outputs_level1_remitted                           <-
with(stardOutputs[stardOutputs$meetInclusion == 1 & stardOutputs$level1_ivra_entered == 1,],
table(level1_remitted, level1_tx, useNA = "always")))

(outputs_level1_remitted_imputed                   <-
with(stardOutputs[stardOutputs$meetInclusion == 1 & stardOutputs$level1_ivra_entered == 1,],
table(level1_remitted_imputed , level1_tx, useNA = "always")))

(outputs_level1_responded                          <-
with(stardOutputs[stardOutputs$meetInclusion == 1 & stardOutputs$level1_ivra_entered == 1,],
table(level1_responded, level1_tx, useNA = "always")))

(outputs_level1_responded_imputed                  <-
with(stardOutputs[stardOutputs$meetInclusion == 1 & stardOutputs$level1_ivra_entered == 1,],
table(level1_responded_imputed , level1_tx, useNA = "always")))

(outputs_level1_fu_ivra_entered                    <-
with(stardOutputs[stardOutputs$meetInclusion == 1 & !(stardOutputs$src_subject_id %in%
c(level2_entered1134 , level2A_entered28, level3_entered313, level4_entered94)),],
table(fu_ivra_entered, level1_tx, useNA = "always")))
```

```
(outputs_level1_fu_relapse                              <-
with(stardOutputs[stardOutputs$meetInclusion == 1 & !(stardOutputs$src_subject_id %in%
c(level2_entered1134 , level2A_entered28, level3_entered313, level4_entered94)),],
table(fu_relapse, level1_tx, useNA = "always")))

(outputs_level1_fu_susRemission_full         <- with(stardOutputs[stardOutputs$meetInclusion == 1
& !(stardOutputs$src_subject_id %in% c(level2_entered1134 , level2A_entered28, level3_entered313,
level4_entered94)),], table(fu_susRemission_full, level1_tx, useNA = "always")))

(outputs_level1_fu_susRemission_weak         <- with(stardOutputs[stardOutputs$meetInclusion == 1
& !(stardOutputs$src_subject_id %in% c(level2_entered1134 , level2A_entered28, level3_entered313,
level4_entered94)),], table(fu_susRemission_weak, level1_tx, useNA = "always")))

(outputs_level1_fu_hasAtLeast1Observations   <- with(stardOutputs[stardOutputs$meetInclusion == 1
& !(stardOutputs$src_subject_id %in% c(level2_entered1134 , level2A_entered28, level3_entered313,
level4_entered94)),], table(fu_hasAtLeast1Observations , level1_tx, useNA = "always")))



(outputs_level1_fu_remissionbeforeFU                           <-
with(stardOutputs[stardOutputs$meetInclusion == 1 & !(stardOutputs$src_subject_id %in%
c(level2_entered1134 , level2A_entered28, level3_entered313, level4_entered94)),],
table(fu_remissionbeforeFU, level1_tx, useNA = "always")))

(outputs_level1_fu_remissionbeforeFU_imputed                   <-
with(stardOutputs[stardOutputs$meetInclusion == 1 & !(stardOutputs$src_subject_id %in%
c(level2_entered1134 , level2A_entered28, level3_entered313, level4_entered94)),],
table(fu_remissionbeforeFU_imputed, level1_tx, useNA = "always")))

(outputs_level1_fu_remitImputed_then_relapse                   <-
with(stardOutputs[stardOutputs$meetInclusion == 1 & !(stardOutputs$src_subject_id %in%
c(level2_entered1134 , level2A_entered28, level3_entered313, level4_entered94)),],
table(fu_remitImputed_then_relapse, level1_tx, useNA = "always")))

(outputs_level1_fu_NOremitImputed_then_relapse                 <-
with(stardOutputs[stardOutputs$meetInclusion == 1 & !(stardOutputs$src_subject_id %in%
c(level2_entered1134 , level2A_entered28, level3_entered313, level4_entered94)),],
table(fu_NOremitImputed_then_relapse , level1_tx, useNA = "always")))

(outputs_level1_fu_remitImputed_then_susRemission_full        <-
with(stardOutputs[stardOutputs$meetInclusion == 1 & !(stardOutputs$src_subject_id %in%
c(level2_entered1134 , level2A_entered28, level3_entered313, level4_entered94)),],
table(fu_remitImputed_then_susRemission_full , level1_tx, useNA = "always")))

(outputs_level1_fu_NOremitImputed_then_susRemission_full      <-
with(stardOutputs[stardOutputs$meetInclusion == 1 & !(stardOutputs$src_subject_id %in%
c(level2_entered1134 , level2A_entered28, level3_entered313, level4_entered94)),],
table(fu_NOremitImputed_then_susRemission_full, level1_tx, useNA = "always")))

(outputs_level1_fu_remitImputed_then_susRemission_weak        <-
with(stardOutputs[stardOutputs$meetInclusion == 1 & !(stardOutputs$src_subject_id %in%
c(level2_entered1134 , level2A_entered28, level3_entered313, level4_entered94)),],
table(fu_remitImputed_then_susRemission_weak, level1_tx, useNA = "always")))

(outputs_level1_fu_NOremitImputed_then_susRemission_weak      <-
with(stardOutputs[stardOutputs$meetInclusion == 1 & !(stardOutputs$src_subject_id %in%
c(level2_entered1134 , level2A_entered28, level3_entered313, level4_entered94)),],
table(fu_NOremitImputed_then_susRemission_weak, level1_tx, useNA = "always")))




## Level 2     ##
```

```
(outputs_level2_count                                <-
with(stardOutputs[stardOutputs$meetInclusion == 1 & (is.na(stardOutputs$level1_remitted ) |
stardOutputs$level1_remitted != 1),] , table(level2_tx, useNA = "always")))

(outputs_level2_remitted                             <-
with(stardOutputs[stardOutputs$meetInclusion == 1 & (is.na(stardOutputs$level1_remitted ) |
stardOutputs$level1_remitted != 1) & stardOutputs$level2_ivra_entered == 1,],
table(level2_remitted, level2_tx, useNA = "always")))

(outputs_level2_remitted_imputed                     <-
with(stardOutputs[stardOutputs$meetInclusion == 1 & (is.na(stardOutputs$level1_remitted ) |
stardOutputs$level1_remitted != 1) & stardOutputs$level2_ivra_entered == 1,],
table(level2_remitted_imputed, level2_tx, useNA = "always")))

(outputs_level2_responded                            <-
with(stardOutputs[stardOutputs$meetInclusion == 1 & (is.na(stardOutputs$level1_remitted ) |
stardOutputs$level1_remitted != 1) & stardOutputs$level2_ivra_entered == 1,],
table(level2_responded, level2_tx, useNA = "always")))

(outputs_level2_responded_imputed                    <-
with(stardOutputs[stardOutputs$meetInclusion == 1 & (is.na(stardOutputs$level1_remitted ) |
stardOutputs$level1_remitted != 1) & stardOutputs$level2_ivra_entered == 1,],
table(level2_responded_imputed, level2_tx, useNA = "always")))

(outputs_level2_fu_ivra_entered                      <-
with(stardOutputs[stardOutputs$src_subject_id %in% level2_entered1134 &
!(stardOutputs$src_subject_id %in% c(level2A_entered28, level3_entered313, level4_entered94)),],
table(fu_ivra_entered, level2_tx, useNA = "always")))

(outputs_level2_fu_relapse                           <-
with(stardOutputs[stardOutputs$src_subject_id %in% level2_entered1134 &
!(stardOutputs$src_subject_id %in% c(level2A_entered28, level3_entered313, level4_entered94)),],
table(fu_relapse, level2_tx, useNA = "always")))

(outputs_level2_fu_susRemission_full         <- with(stardOutputs[stardOutputs$src_subject_id
%in% level2_entered1134 & !(stardOutputs$src_subject_id %in% c(level2A_entered28,
level3_entered313, level4_entered94)),], table(fu_susRemission_full, level2_tx, useNA =
"always")))

(outputs_level2_fu_susRemission_weak         <- with(stardOutputs[stardOutputs$src_subject_id
%in% level2_entered1134 & !(stardOutputs$src_subject_id %in% c(level2A_entered28,
level3_entered313, level4_entered94)),], table(fu_susRemission_weak, level2_tx, useNA =
"always")))

(outputs_level2_fu_hasAtLeast1Observations   <- with(stardOutputs[stardOutputs$src_subject_id
%in% level2_entered1134 & !(stardOutputs$src_subject_id %in% c(level2A_entered28,
level3_entered313, level4_entered94)),], table(fu_hasAtLeast1Observations , level2_tx, useNA =
"always")))



(outputs_level2_fu_remissionbeforeFU                         <-
with(stardOutputs[stardOutputs$src_subject_id %in% level2_entered1134 &
!(stardOutputs$src_subject_id %in% c(level2A_entered28, level3_entered313, level4_entered94)),],
table(fu_remissionbeforeFU, level2_tx, useNA = "always")))

(outputs_level2_fu_remissionbeforeFU_imputed                 <-
with(stardOutputs[stardOutputs$src_subject_id %in% level2_entered1134 &
!(stardOutputs$src_subject_id %in% c(level2A_entered28, level3_entered313, level4_entered94)),],
table(fu_remissionbeforeFU_imputed, level2_tx, useNA = "always")))

(outputs_level2_fu_remitImputed_then_relapse                 <-
with(stardOutputs[stardOutputs$src_subject_id %in% level2_entered1134 &
!(stardOutputs$src_subject_id %in% c(level2A_entered28, level3_entered313, level4_entered94)),],
table(fu_remitImputed_then_relapse, level2_tx, useNA = "always")))

(outputs_level2_fu_NOremitImputed_then_relapse               <-
with(stardOutputs[stardOutputs$src_subject_id %in% level2_entered1134 &
```

```
!(stardOutputs$src_subject_id %in% c(level2A_entered28, level3_entered313, level4_entered94)),],
table(fu_NOremitImputed_then_relapse , level2_tx, useNA = "always")))

(outputs_level2_fu_remitImputed_then_susRemission_full      <-
with(stardOutputs[stardOutputs$src_subject_id %in% level2_entered1134 &
!(stardOutputs$src_subject_id %in% c(level2A_entered28, level3_entered313, level4_entered94)),],
table(fu_remitImputed_then_susRemission_full , level2_tx, useNA = "always")))

(outputs_level2_fu_NOremitImputed_then_susRemission_full    <-
with(stardOutputs[stardOutputs$src_subject_id %in% level2_entered1134 &
!(stardOutputs$src_subject_id %in% c(level2A_entered28, level3_entered313, level4_entered94)),],
table(fu_NOremitImputed_then_susRemission_full, level2_tx, useNA = "always")))

(outputs_level2_fu_remitImputed_then_susRemission_weak      <-
with(stardOutputs[stardOutputs$src_subject_id %in% level2_entered1134 &
!(stardOutputs$src_subject_id %in% c(level2A_entered28, level3_entered313, level4_entered94)),],
table(fu_remitImputed_then_susRemission_weak, level2_tx, useNA = "always")))

(outputs_level2_fu_NOremitImputed_then_susRemission_weak    <-
with(stardOutputs[stardOutputs$src_subject_id %in% level2_entered1134 &
!(stardOutputs$src_subject_id %in% c(level2A_entered28, level3_entered313, level4_entered94)),],
table(fu_NOremitImputed_then_susRemission_weak, level2_tx, useNA = "always")))




## Level 2A    ##

(outputs_level2A_count                              <-
with(stardOutputs[stardOutputs$meetInclusion == 1 & (is.na(stardOutputs$level1_remitted ) |
stardOutputs$level1_remitted != 1) & (is.na(stardOutputs$level2_remitted ) |
stardOutputs$level2_remitted != 1) & stardOutputs$level2A_ivra_entered == 1,], table(level2A_tx,
useNA = "always")))

(outputs_level2A_remitted                           <-
with(stardOutputs[stardOutputs$meetInclusion == 1 & (is.na(stardOutputs$level1_remitted ) |
stardOutputs$level1_remitted != 1) & (is.na(stardOutputs$level2_remitted ) |
stardOutputs$level2_remitted != 1) & stardOutputs$level2A_ivra_entered == 1,],
table(level2A_remitted, level2A_tx, useNA = "always")))

(outputs_level2A_remitted_imputed                   <-
with(stardOutputs[stardOutputs$meetInclusion == 1 & (is.na(stardOutputs$level1_remitted ) |
stardOutputs$level1_remitted != 1) & (is.na(stardOutputs$level2_remitted ) |
stardOutputs$level2_remitted != 1) & stardOutputs$level2A_ivra_entered == 1,],
table(level2A_remitted_imputed, level2A_tx, useNA = "always")))

(outputs_level2A_responded                          <-
with(stardOutputs[stardOutputs$meetInclusion == 1 & (is.na(stardOutputs$level1_remitted ) |
stardOutputs$level1_remitted != 1) & (is.na(stardOutputs$level2_remitted ) |
stardOutputs$level2_remitted != 1) & stardOutputs$level2A_ivra_entered == 1,],
table(level2A_responded, level2A_tx, useNA = "always")))

(outputs_level2A_responded_imputed                  <-
with(stardOutputs[stardOutputs$meetInclusion == 1 & (is.na(stardOutputs$level1_remitted ) |
stardOutputs$level1_remitted != 1) & (is.na(stardOutputs$level2_remitted ) |
```

```
stardOutputs$level2_remitted != 1) & stardOutputs$level2A_ivra_entered == 1,],
table(level2A_responded_imputed, level2A_tx, useNA = "always")))

(outputs_level2A_fu_ivra_entered                    <-
with(stardOutputs[stardOutputs$src_subject_id %in% level2A_entered28 &
!(stardOutputs$src_subject_id %in% c(level3_entered313, 13, level4_entered94)),],
table(fu_ivra_entered, level2A_tx, useNA = "always")))

(outputs_level2A_fu_relapse                         <-
with(stardOutputs[stardOutputs$src_subject_id %in% level2A_entered28 &
!(stardOutputs$src_subject_id %in% c(level3_entered313, 13, level4_entered94)),],
table(fu_relapse, level2A_tx, useNA = "always")))

(outputs_level2A_fu_susRemission_full        <- with(stardOutputs[stardOutputs$src_subject_id
%in% level2A_entered28 & !(stardOutputs$src_subject_id %in% c(level3_entered313, 13,
level4_entered94)),], table(fu_susRemission_full, level2A_tx, useNA = "always")))

(outputs_level2A_fu_susRemission_weak        <- with(stardOutputs[stardOutputs$src_subject_id
%in% level2A_entered28 & !(stardOutputs$src_subject_id %in% c(level3_entered313, 13,
level4_entered94)),], table(fu_susRemission_weak, level2A_tx, useNA = "always")))

(outputs_level2A_fu_hasAtLeast1Observations  <- with(stardOutputs[stardOutputs$src_subject_id
%in% level2A_entered28 & !(stardOutputs$src_subject_id %in% c(level3_entered313, 13,
level4_entered94)),], table(fu_hasAtLeast1Observations , level2A_tx, useNA = "always")))




(outputs_level2A_fu_remissionbeforeFU               <-
with(stardOutputs[stardOutputs$src_subject_id %in% level2A_entered28 &
!(stardOutputs$src_subject_id %in% c(level3_entered313, 13, level4_entered94)),],
table(fu_remissionbeforeFU, level2A_tx, useNA = "always")))

(outputs_level2A_fu_remissionbeforeFU_imputed               <-
with(stardOutputs[stardOutputs$src_subject_id %in% level2A_entered28 &
!(stardOutputs$src_subject_id %in% c(level3_entered313, 13, level4_entered94)),],
table(fu_remissionbeforeFU_imputed, level2A_tx, useNA = "always")))

(outputs_level2A_fu_remitImputed_then_relapse                      <-
with(stardOutputs[stardOutputs$src_subject_id %in% level2A_entered28 &
!(stardOutputs$src_subject_id %in% c(level3_entered313, 13, level4_entered94)),],
table(fu_remitImputed_then_relapse, level2A_tx, useNA = "always")))

(outputs_level2A_fu_NOremitImputed_then_relapse             <-
with(stardOutputs[stardOutputs$src_subject_id %in% level2A_entered28 &
!(stardOutputs$src_subject_id %in% c(level3_entered313, 13, level4_entered94)),],
table(fu_NOremitImputed_then_relapse , level2A_tx, useNA = "always")))

(outputs_level2A_fu_remitImputed_then_susRemission_full     <-
with(stardOutputs[stardOutputs$src_subject_id %in% level2A_entered28 &
!(stardOutputs$src_subject_id %in% c(level3_entered313, 13, level4_entered94)),],
table(fu_remitImputed_then_susRemission_full , level2A_tx, useNA = "always")))

(outputs_level2A_fu_NOremitImputed_then_susRemission_full   <-
with(stardOutputs[stardOutputs$src_subject_id %in% level2A_entered28 &
!(stardOutputs$src_subject_id %in% c(level3_entered313, 13, level4_entered94)),],
table(fu_NOremitImputed_then_susRemission_full, level2A_tx, useNA = "always")))

(outputs_level2A_fu_remitImputed_then_susRemission_weak     <-
with(stardOutputs[stardOutputs$src_subject_id %in% level2A_entered28 &
!(stardOutputs$src_subject_id %in% c(level3_entered313, 13, level4_entered94)),],
table(fu_remitImputed_then_susRemission_weak, level2A_tx, useNA = "always")))

(outputs_level2A_fu_NOremitImputed_then_susRemission_weak   <-
with(stardOutputs[stardOutputs$src_subject_id %in% level2A_entered28 &
!(stardOutputs$src_subject_id %in% c(level3_entered313, 13, level4_entered94)),],
table(fu_NOremitImputed_then_susRemission_weak, level2A_tx, useNA = "always")))
```

```
## Level 3     ##

(outputs_level3_count                              <-
with(stardOutputs[stardOutputs$meetInclusion == 1 & (is.na(stardOutputs$level1_remitted ) |
stardOutputs$level1_remitted != 1) & (is.na(stardOutputs$level2_remitted ) |
stardOutputs$level2_remitted != 1) & (is.na(stardOutputs$level2A_remitted ) |
stardOutputs$level2A_remitted != 1)  & stardOutputs$level3_ivra_entered == 1,], table(level3_tx,
useNA = "always")))

(outputs_level3_remitted                           <-
with(stardOutputs[stardOutputs$meetInclusion == 1  & (is.na(stardOutputs$level1_remitted ) |
stardOutputs$level1_remitted != 1) & (is.na(stardOutputs$level2_remitted ) |
stardOutputs$level2_remitted != 1) & (is.na(stardOutputs$level2A_remitted ) |
stardOutputs$level2A_remitted != 1)  & stardOutputs$level3_ivra_entered == 1,],
table(level3_remitted, level3_tx, useNA = "always")))

(outputs_level3_remitted_imputed                   <-
with(stardOutputs[stardOutputs$meetInclusion == 1  & (is.na(stardOutputs$level1_remitted ) |
stardOutputs$level1_remitted != 1) & (is.na(stardOutputs$level2_remitted ) |
stardOutputs$level2_remitted != 1) & (is.na(stardOutputs$level2A_remitted ) |
stardOutputs$level2A_remitted != 1)  & stardOutputs$level3_ivra_entered == 1,],
table(level3_remitted_imputed, level3_tx, useNA = "always")))

(outputs_level3_responded                          <-
with(stardOutputs[stardOutputs$meetInclusion == 1  & (is.na(stardOutputs$level1_remitted ) |
stardOutputs$level1_remitted != 1) & (is.na(stardOutputs$level2_remitted ) |
stardOutputs$level2_remitted != 1) & (is.na(stardOutputs$level2A_remitted ) |
stardOutputs$level2A_remitted != 1)  & stardOutputs$level3_ivra_entered == 1,],
table(level3_responded, level3_tx, useNA = "always")))

(outputs_level3_responded_imputed                  <-
with(stardOutputs[stardOutputs$meetInclusion == 1  & (is.na(stardOutputs$level1_remitted ) |
stardOutputs$level1_remitted != 1) & (is.na(stardOutputs$level2_remitted ) |
stardOutputs$level2_remitted != 1) & (is.na(stardOutputs$level2A_remitted ) |
stardOutputs$level2A_remitted != 1)  & stardOutputs$level3_ivra_entered == 1,],
table(level3_responded_imputed, level3_tx, useNA = "always")))

(outputs_level3_fu_ivra_entered                    <-
with(stardOutputs[stardOutputs$src_subject_id %in% level3_entered313 &
!(stardOutputs$src_subject_id %in% c(level4_entered94)),], table(fu_ivra_entered, level3_tx,
useNA = "always")))

(outputs_level3_fu_relapse                         <-
with(stardOutputs[stardOutputs$src_subject_id %in% level3_entered313 &
!(stardOutputs$src_subject_id %in% c(level4_entered94)),], table(fu_relapse, level3_tx, useNA =
"always")))

(outputs_level3_fu_susRemission_full         <- with(stardOutputs[stardOutputs$src_subject_id
%in% level3_entered313 & !(stardOutputs$src_subject_id %in% c(level4_entered94)),],
table(fu_susRemission_full, level3_tx, useNA = "always")))

(outputs_level3_fu_susRemission_weak         <- with(stardOutputs[stardOutputs$src_subject_id
%in% level3_entered313 & !(stardOutputs$src_subject_id %in% c(level4_entered94)),],
table(fu_susRemission_weak, level3_tx, useNA = "always")))

(outputs_level3_fu_hasAtLeast1Observations   <- with(stardOutputs[stardOutputs$src_subject_id
%in% level3_entered313 & !(stardOutputs$src_subject_id %in% c(level4_entered94)),],
table(fu_hasAtLeast1Observations , level3_tx, useNA = "always")))



(outputs_level3_fu_remissionbeforeFU               <-
with(stardOutputs[stardOutputs$src_subject_id %in% level3_entered313 &
```

```
!(stardOutputs$src_subject_id %in% c(level4_entered94)),], table(fu_remissionbeforeFU, level3_tx, useNA = "always")))

(outputs_level3_fu_remissionbeforeFU_imputed        <- with(stardOutputs[stardOutputs$src_subject_id %in% level3_entered313 & !(stardOutputs$src_subject_id %in% c(level4_entered94)),], table(fu_remissionbeforeFU_imputed, level3_tx, useNA = "always")))

(outputs_level3_fu_remitImputed_then_relapse               <- with(stardOutputs[stardOutputs$src_subject_id %in% level3_entered313 & !(stardOutputs$src_subject_id %in% c(level4_entered94)),], table(fu_remitImputed_then_relapse, level3_tx, useNA = "always")))

(outputs_level3_fu_NOremitImputed_then_relapse             <- with(stardOutputs[stardOutputs$src_subject_id %in% level3_entered313 & !(stardOutputs$src_subject_id %in% c(level4_entered94)),], table(fu_NOremitImputed_then_relapse , level3_tx, useNA = "always")))

(outputs_level3_fu_remitImputed_then_susRemission_full      <- with(stardOutputs[stardOutputs$src_subject_id %in% level3_entered313 & !(stardOutputs$src_subject_id %in% c(level4_entered94)),], table(fu_remitImputed_then_susRemission_full , level3_tx, useNA = "always")))

(outputs_level3_fu_NOremitImputed_then_susRemission_full    <- with(stardOutputs[stardOutputs$src_subject_id %in% level3_entered313 & !(stardOutputs$src_subject_id %in% c(level4_entered94)),], table(fu_NOremitImputed_then_susRemission_full, level3_tx, useNA = "always")))

(outputs_level3_fu_remitImputed_then_susRemission_weak      <- with(stardOutputs[stardOutputs$src_subject_id %in% level3_entered313 & !(stardOutputs$src_subject_id %in% c(level4_entered94)),], table(fu_remitImputed_then_susRemission_weak, level3_tx, useNA = "always")))

(outputs_level3_fu_NOremitImputed_then_susRemission_weak    <- with(stardOutputs[stardOutputs$src_subject_id %in% level3_entered313 & !(stardOutputs$src_subject_id %in% c(level4_entered94)),], table(fu_NOremitImputed_then_susRemission_weak, level3_tx, useNA = "always")))




## Level 4     ##

(outputs_level4_count                               <- with(stardOutputs[stardOutputs$meetInclusion == 1 & (is.na(stardOutputs$level1_remitted ) | stardOutputs$level1_remitted != 1) & (is.na(stardOutputs$level2_remitted ) | stardOutputs$level2_remitted != 1) & (is.na(stardOutputs$level2A_remitted ) | stardOutputs$level2A_remitted != 1)  & (is.na(stardOutputs$level3_remitted ) | stardOutputs$level3_remitted != 1)  & stardOutputs$level4_ivra_entered == 1,], table(level4_tx, useNA = "always")))

(outputs_level4_remitted                            <- with(stardOutputs[stardOutputs$meetInclusion == 1 & (is.na(stardOutputs$level1_remitted ) | stardOutputs$level1_remitted != 1) & (is.na(stardOutputs$level2_remitted ) | stardOutputs$level2_remitted != 1) & (is.na(stardOutputs$level2A_remitted ) | stardOutputs$level2A_remitted != 1)  & (is.na(stardOutputs$level3_remitted ) | stardOutputs$level3_remitted != 1)  & stardOutputs$level4_ivra_entered == 1,], table(level4_remitted, level4_tx, useNA = "always")))

(outputs_level4_remitted_imputed                    <- with(stardOutputs[stardOutputs$meetInclusion == 1 & (is.na(stardOutputs$level1_remitted ) | stardOutputs$level1_remitted != 1) & (is.na(stardOutputs$level2_remitted ) | stardOutputs$level2_remitted != 1) & (is.na(stardOutputs$level2A_remitted ) | stardOutputs$level2A_remitted != 1)  & (is.na(stardOutputs$level3_remitted ) | stardOutputs$level3_remitted != 1)  & stardOutputs$level4_ivra_entered == 1,], table(level4_remitted_imputed, level4_tx, useNA = "always")))
```

```
(outputs_level4_responded                           <-
with(stardOutputs[stardOutputs$meetInclusion == 1 & (is.na(stardOutputs$level1_remitted ) |
stardOutputs$level1_remitted != 1) & (is.na(stardOutputs$level2_remitted ) |
stardOutputs$level2_remitted != 1) & (is.na(stardOutputs$level2A_remitted ) |
stardOutputs$level2A_remitted != 1)  & (is.na(stardOutputs$level3_remitted ) |
stardOutputs$level3_remitted != 1)  & stardOutputs$level4_ivra_entered == 1,],
table(level4_responded, level4_tx, useNA = "always")))

(outputs_level4_responded_imputed                   <-
with(stardOutputs[stardOutputs$meetInclusion == 1 & (is.na(stardOutputs$level1_remitted ) |
stardOutputs$level1_remitted != 1) & (is.na(stardOutputs$level2_remitted ) |
stardOutputs$level2_remitted != 1) & (is.na(stardOutputs$level2A_remitted ) |
stardOutputs$level2A_remitted != 1)  & (is.na(stardOutputs$level3_remitted ) |
stardOutputs$level3_remitted != 1)  & stardOutputs$level4_ivra_entered == 1,],
table(level4_responded_imputed, level4_tx, useNA = "always")))

(outputs_level4_fu_ivra_entered                     <-
with(stardOutputs[stardOutputs$src_subject_id %in% level4_entered94,], table(fu_ivra_entered,
level4_tx, useNA = "always")))

(outputs_level4_fu_relapse                          <-
with(stardOutputs[stardOutputs$src_subject_id %in% level4_entered94,], table(fu_relapse,
level4_tx, useNA = "always")))

(outputs_level4_fu_susRemission_full         <- with(stardOutputs[stardOutputs$src_subject_id
%in% level4_entered94,], table(fu_susRemission_full, level4_tx, useNA = "always")))

(outputs_level4_fu_susRemission_weak         <- with(stardOutputs[stardOutputs$src_subject_id
%in% level4_entered94,], table(fu_susRemission_weak, level4_tx, useNA = "always")))

(outputs_level4_fu_hasAtLeast1Observations          <-
with(stardOutputs[stardOutputs$src_subject_id %in% level4_entered94,],
table(fu_hasAtLeast1Observations , level4_tx, useNA = "always")))




(outputs_level4_fu_remissionbeforeFU                <-
with(stardOutputs[stardOutputs$src_subject_id %in% level4_entered94,],
table(fu_remissionbeforeFU, level4_tx, useNA = "always")))

(outputs_level4_fu_remissionbeforeFU_imputed        <-
with(stardOutputs[stardOutputs$src_subject_id %in% level4_entered94,],
table(fu_remissionbeforeFU_imputed, level4_tx, useNA = "always")))



(outputs_level4_fu_remitImputed_then_relapse                <-
with(stardOutputs[stardOutputs$src_subject_id %in% level4_entered94,],
table(fu_remitImputed_then_relapse, level4_tx, useNA = "always")))

(outputs_level4_fu_NOremitImputed_then_relapse              <-
with(stardOutputs[stardOutputs$src_subject_id %in% level4_entered94,],
table(fu_NOremitImputed_then_relapse , level4_tx, useNA = "always")))

(outputs_level4_fu_remitImputed_then_susRemission_full      <-
with(stardOutputs[stardOutputs$src_subject_id %in% level4_entered94,],
table(fu_remitImputed_then_susRemission_full , level4_tx, useNA = "always")))

(outputs_level4_fu_NOremitImputed_then_susRemission_full    <-
with(stardOutputs[stardOutputs$src_subject_id %in% level4_entered94,],
table(fu_NOremitImputed_then_susRemission_full, level4_tx, useNA = "always")))

(outputs_level4_fu_remitImputed_then_susRemission_weak      <-
with(stardOutputs[stardOutputs$src_subject_id %in% level4_entered94,],
table(fu_remitImputed_then_susRemission_weak, level4_tx, useNA = "always")))
```

```
(outputs_level4_fu_NOremitImputed_then_susRemission_weak    <-
with(stardOutputs[stardOutputs$src_subject_id %in% level4_entered94,],
table(fu_NOremitImputed_then_susRemission_weak, level4_tx, useNA = "always")))
```

```
## Function to extract attributes from output table  ##

extract <- function(outputTable){

        rownames(outputTable) <- paste(names(attributes(outputTable)$dimnames)[[1]],
rownames(outputTable), sep = " ")

        return(outputTable)

}



### Merging and savings outputs       ###



summaryLevel1 <- rbind(outputs_level1_count, extract( outputs_level1_remitted),
extract(outputs_level1_remitted_imputed), extract( outputs_level1_responded),
extract(outputs_level1_responded_imputed), extract(outputs_level1_fu_ivra_entered),  extract(
outputs_level1_fu_remissionbeforeFU), extract(
outputs_level1_fu_remissionbeforeFU_imputed),extract( outputs_level1_fu_relapse), extract(
outputs_level1_fu_susRemission_full), extract(outputs_level1_fu_susRemission_weak),
extract(outputs_level1_fu_hasAtLeast1Observations),
extract(outputs_level1_fu_remitImputed_then_relapse),extract(outputs_level1_fu_NOremitImputed_the
n_relapse), extract(outputs_level1_fu_remitImputed_then_susRemission_full),
extract(outputs_level1_fu_NOremitImputed_then_susRemission_full),
        extract(outputs_level1_fu_remitImputed_then_susRemission_weak),
        extract(outputs_level1_fu_NOremitImputed_then_susRemission_weak))

summaryLevel2 <- rbind(outputs_level2_count, extract( outputs_level2_remitted),
extract(outputs_level2_remitted_imputed), extract( outputs_level2_responded),
extract(outputs_level2_responded_imputed), extract(outputs_level2_fu_ivra_entered),  extract(
outputs_level2_fu_remissionbeforeFU), extract(
outputs_level2_fu_remissionbeforeFU_imputed),extract( outputs_level2_fu_relapse), extract(
outputs_level2_fu_susRemission_full), extract(outputs_level2_fu_susRemission_weak),
extract(outputs_level2_fu_hasAtLeast1Observations),
extract(outputs_level2_fu_remitImputed_then_relapse),extract(outputs_level2_fu_NOremitImputed_the
n_relapse), extract(outputs_level2_fu_remitImputed_then_susRemission_full),
extract(outputs_level2_fu_NOremitImputed_then_susRemission_full),
        extract(outputs_level2_fu_remitImputed_then_susRemission_weak),
        extract(outputs_level2_fu_NOremitImputed_then_susRemission_weak))

summaryLevel2A <- rbind(outputs_level2A_count, extract( outputs_level2A_remitted),
extract(outputs_level2A_remitted_imputed), extract( outputs_level2A_responded),
extract(outputs_level2A_responded_imputed), extract(outputs_level2A_fu_ivra_entered),  extract(
outputs_level2A_fu_remissionbeforeFU), extract(
outputs_level2A_fu_remissionbeforeFU_imputed),extract( outputs_level2A_fu_relapse), extract(
outputs_level2A_fu_susRemission_full), extract(outputs_level2A_fu_susRemission_weak),
extract(outputs_level2A_fu_hasAtLeast1Observations),
extract(outputs_level2A_fu_remitImputed_then_relapse),extract(outputs_level2A_fu_NOremitImputed_t
hen_relapse), extract(outputs_level2A_fu_remitImputed_then_susRemission_full),
extract(outputs_level2A_fu_NOremitImputed_then_susRemission_full),
        extract(outputs_level2A_fu_remitImputed_then_susRemission_weak),
        extract(outputs_level2A_fu_NOremitImputed_then_susRemission_weak))
```

```
summaryLevel3 <- rbind(outputs_level3_count, extract( outputs_level3_remitted),
extract(outputs_level3_remitted_imputed), extract( outputs_level3_responded),
extract(outputs_level3_responded_imputed), extract(outputs_level3_fu_ivra_entered),  extract(
outputs_level3_fu_remissionbeforeFU), extract(
outputs_level3_fu_remissionbeforeFU_imputed),extract( outputs_level3_fu_relapse), extract(
outputs_level3_fu_susRemission_full), extract(outputs_level3_fu_susRemission_weak),
extract(outputs_level3_fu_hasAtLeast1Observations),
extract(outputs_level3_fu_remitImputed_then_relapse),extract(outputs_level3_fu_NOremitImputed_the
n_relapse), extract(outputs_level3_fu_remitImputed_then_susRemission_full),
extract(outputs_level3_fu_NOremitImputed_then_susRemission_full),
       extract(outputs_level3_fu_remitImputed_then_susRemission_weak),
       extract(outputs_level3_fu_NOremitImputed_then_susRemission_weak))

summaryLevel4 <- rbind(outputs_level4_count, extract( outputs_level4_remitted),
extract(outputs_level4_remitted_imputed), extract( outputs_level4_responded),
extract(outputs_level4_responded_imputed), extract(outputs_level4_fu_ivra_entered),  extract(
outputs_level4_fu_remissionbeforeFU), extract(
outputs_level4_fu_remissionbeforeFU_imputed),extract( outputs_level4_fu_relapse), extract(
outputs_level4_fu_susRemission_full), extract(outputs_level4_fu_susRemission_weak),
extract(outputs_level4_fu_hasAtLeast1Observations),
extract(outputs_level4_fu_remitImputed_then_relapse),extract(outputs_level4_fu_NOremitImputed_the
n_relapse), extract(outputs_level4_fu_remitImputed_then_susRemission_full),
extract(outputs_level4_fu_NOremitImputed_then_susRemission_full),
       extract(outputs_level4_fu_remitImputed_then_susRemission_weak),
       extract(outputs_level4_fu_NOremitImputed_then_susRemission_weak))
```

```
### Outputting HRSD Change
```

```
hrsdChangeImputedLevel1 <- with(stardOutputs[stardOutputs$meetInclusion == 1 &
stardOutputs$level1_ivra_entered == 1,], aggregate(level1_hrsdchange_imputed, list(level1_tx),
summarizeOutputs))

hrsdChangeImputedLevel2 <- with(stardOutputs[stardOutputs$meetInclusion == 1 &
(is.na(stardOutputs$level1_remitted ) | stardOutputs$level1_remitted != 1) &
stardOutputs$level2_ivra_entered == 1,], aggregate(level2_hrsdchange_imputed, list(level2_tx),
summarizeOutputs))

hrsdChangeImputedLevel2A <- with(stardOutputs[stardOutputs$meetInclusion == 1 &
(is.na(stardOutputs$level1_remitted ) | stardOutputs$level1_remitted != 1) &
(is.na(stardOutputs$level2_remitted ) | stardOutputs$level2_remitted != 1) &
stardOutputs$level2A_ivra_entered == 1,], aggregate(level2A_hrsdchange_imputed, list(level2A_tx),
summarizeOutputs))

hrsdChangeImputedLevel3 <- with(stardOutputs[stardOutputs$meetInclusion == 1  &
(is.na(stardOutputs$level1_remitted ) | stardOutputs$level1_remitted != 1) &
(is.na(stardOutputs$level2_remitted ) | stardOutputs$level2_remitted != 1) &
(is.na(stardOutputs$level2A_remitted ) | stardOutputs$level2A_remitted != 1)  &
stardOutputs$level3_ivra_entered == 1,], aggregate(level3_hrsdchange_imputed, list(level3_tx),
summarizeOutputs))

hrsdChangeImputedLevel4 <- with(stardOutputs[stardOutputs$meetInclusion == 1 &
(is.na(stardOutputs$level1_remitted ) | stardOutputs$level1_remitted != 1) &
(is.na(stardOutputs$level2_remitted ) | stardOutputs$level2_remitted != 1) &
(is.na(stardOutputs$level2A_remitted ) | stardOutputs$level2A_remitted != 1)  &
(is.na(stardOutputs$level3_remitted ) | stardOutputs$level3_remitted != 1)  &
stardOutputs$level4_ivra_entered == 1,], aggregate(level4_hrsdchange_imputed, list(level4_tx),
summarizeOutputs))
```

```
hrsdChangenonImputedLevel1 <- with(stardOutputs[stardOutputs$meetInclusion == 1 &
stardOutputs$level1_ivra_entered == 1,], aggregate(level1_hrsdchange, list(level1_tx),
summarizeOutputs))

hrsdChangenonImputedLevel2 <- with(stardOutputs[stardOutputs$meetInclusion == 1 &
(is.na(stardOutputs$level1_remitted ) | stardOutputs$level1_remitted != 1) &
stardOutputs$level2_ivra_entered == 1,], aggregate(level2_hrsdchange, list(level2_tx),
summarizeOutputs))

hrsdChangenonImputedLevel2A <- with(stardOutputs[stardOutputs$meetInclusion == 1 &
(is.na(stardOutputs$level1_remitted ) | stardOutputs$level1_remitted != 1) &
(is.na(stardOutputs$level2_remitted ) | stardOutputs$level2_remitted != 1) &
stardOutputs$level2A_ivra_entered == 1,], aggregate(level2A_hrsdchange, list(level2A_tx),
summarizeOutputs))

hrsdChangenonImputedLevel3 <- with(stardOutputs[stardOutputs$meetInclusion == 1  &
(is.na(stardOutputs$level1_remitted ) | stardOutputs$level1_remitted != 1) &
(is.na(stardOutputs$level2_remitted ) | stardOutputs$level2_remitted != 1) &
(is.na(stardOutputs$level2A_remitted ) | stardOutputs$level2A_remitted != 1)  &
stardOutputs$level3_ivra_entered == 1,], aggregate(level3_hrsdchange, list(level3_tx),
summarizeOutputs))

hrsdChangenonImputedLevel4 <- with(stardOutputs[stardOutputs$meetInclusion == 1 &
(is.na(stardOutputs$level1_remitted ) | stardOutputs$level1_remitted != 1) &
(is.na(stardOutputs$level2_remitted ) | stardOutputs$level2_remitted != 1) &
(is.na(stardOutputs$level2A_remitted ) | stardOutputs$level2A_remitted != 1)  &
(is.na(stardOutputs$level3_remitted ) | stardOutputs$level3_remitted != 1)  &
stardOutputs$level4_ivra_entered == 1,], aggregate(level4_hrsdchange, list(level4_tx),
summarizeOutputs))



write.csv(t(summaryLevel1), "STARD Summary Level1 - new NIMH.csv")

write.csv(t(summaryLevel2), "STARD Summary Level2 - new NIMH.csv")

write.csv(t(summaryLevel2A), "STARD Summary Level2A - new NIMH.csv")

write.csv(t(summaryLevel3), "STARD Summary Level3 - new NIMH.csv")

write.csv(t(summaryLevel4), "STARD Summary Level4 - new NIMH.csv")


write.csv(t(hrsdChangeImputedLevel1 ), "STARD hrsdChange_Imputed Level1 - new NIMH.csv")

write.csv(t(hrsdChangeImputedLevel2 ), "STARD hrsdChange_Imputed Level2 - new NIMH.csv")

write.csv(t(hrsdChangeImputedLevel2A ), "STARD hrsdChange_Imputed Level2A - new NIMH.csv")

write.csv(t(hrsdChangeImputedLevel3 ), "STARD hrsdChange_Imputed Level3 - new NIMH.csv")

write.csv(t(hrsdChangeImputedLevel4 ), "STARD hrsdChange_Imputed Level4 - new NIMH.csv")



write.csv(t(hrsdChangenonImputedLevel1 ), "STARD hrsdChange_nonImputed Level1 - new NIMH.csv")

write.csv(t(hrsdChangenonImputedLevel2 ), "STARD hrsdChange_nonImputed Level2 - new NIMH.csv")

write.csv(t(hrsdChangenonImputedLevel2A ), "STARD hrsdChange_nonImputed Level2A - new NIMH.csv")

write.csv(t(hrsdChangenonImputedLevel3 ), "STARD hrsdChange_nonImputed Level3 - new NIMH.csv")
```

```
write.csv(t(hrsdChangenonImputedLevel4 ), "STARD hrsdChange_nonImputed Level4 - new NIMH.csv")

write.csv(stardOutputs, "STARD - patientOutputs - new NIMH.csv")

stardOutputsNew       <- read.csv("STARD - patientOutputs.csv")

stardOutputsNew $X <- NULL

stardOutputsNew[order(stardOutputsNew$src_subject_id),]  ==
stardOutputs[order(stardOutputs$src_subject_id),]

all.equal(stardOutputsNew[order(stardOutputsNew$src_subject_id),],stardOutputs[order(stardOutputs
$src_subject_id),])

all.equal(stardOutputs[order(stardOutputs$src_subject_id),], stardOutputsNew)

################################  Level 2 Exclusions #####################################
```

```
##### CAREFUL THESE EXCLUSION CATEGOREIS ARE OVERLAPPING

(outputs_level2_exclude_didNotMeetL1Inclusion                              <-
with(stardOutputs[stardOutputs$meetInclusion == 0,], table(level2_tx)))

(outputs_level2_exclude_meetInclusionbutlastLevel1hrsdRemitted             <-
with(stardOutputs[stardOutputs$meetInclusion == 1 & !is.na(stardOutputs$level1_last_hrsd_sum) &
stardOutputs$level1_last_hrsd_sum<= 7,], table(level2_tx)))

(outputs_level2_exclude_lastLevel1hrsdRemitted_doNotDoubleCount            <-
with(stardOutputs[!is.na(stardOutputs$level1_last_hrsd_sum) & stardOutputs$level1_last_hrsd_sum<=
7,], table(level2_tx)))               #Do not use this, it double counts

(outputs_level2_exclude_didNotMeetL1orL2_doNotDoubleCount                  <-
with(stardOutputs[stardOutputs$meetInclusion == 0 & !is.na(stardOutputs$level1_last_hrsd_sum) &
stardOutputs$level1_last_hrsd_sum<= 7,], table(level2_tx))) #Do not use this, it double counts
```

```
### Saving Level 2 Exclusion counts   ###

summaryLevel2_exclusions <-   rbind(

                              nTxLevel2 = outputs_level2_count
[!is.na(names(outputs_level2_count))]   ,

                              exclude_didNotMeetL1Inclusion =
outputs_level2_exclude_didNotMeetL1Inclusion,

                              exclude_meetInclusionbutlastLevel1hrsdRemitted =
outputs_level2_exclude_meetInclusionbutlastLevel1hrsdRemitted,

                              exclude_lastLevel1hrsdRemitted_doNotDoubleCount =
outputs_level2_exclude_lastLevel1hrsdRemitted_doNotDoubleCount,

                              exclude_didNotMeetL1orL2_doNotDoubleCount =
outputs_level2_exclude_didNotMeetL1orL2_doNotDoubleCount)


write.csv(summaryLevel2_exclusions,  "level2 Exclusions.csv")
```

**Supplement 6:**
**Demographics and clinical characteristics**

| | Citalopram plus Bupropion (n=216) | | Citalopram plus Buspirone (n=225) | | Total Augmentation (n=441) | |
|---|---|---|---|---|---|---|
| **Demographic** | **Mean** | **SD** | **Mean** | **SD** | **Mean** | **SD** |
| **Age** | 40.9 | 12.7 | 42.4 | 12.6 | 41.7 | 12.7 |
| **Education (years)** | 13.3 | 3.3 | 12.6 | 3.6 | 12.9 | 3.5 |
| **Monthly household income** | 2121.5 | 2214.7 | 2125.5 | 2887.8 | 2123.5 | 2578.2 |
| | **n** | **%** | **n** | **%** | **n** | **%** |
| **Female** | 138 | 63.9 | 126 | 56.0 | 264 | 59.9 |
| **Race** | | | | | | |
| **White** | 170 | 78.7 | 178 | 79.1 | 348 | 78.9 |
| **Black** | 18 | 8.3 | 19 | 8.4 | 37 | 8.4 |
| **Other** | 28 | 13.0 | 28 | 12.4 | 56 | 12.7 |
| **Hispanic or Latino** | 30 | 13.9 | 33 | 14.7 | 63 | 14.3 |
| **Employment status** | | | | | | |
| **Employed** | 125 | 57.9 | 109 | 48.4 | 234 | 53.1 |
| **Unemployed** | 85 | 39.4 | 105 | 46.7 | 190 | 43.1 |
| **Retired** | 5 | 2.3 | 11 | 4.9 | 16 | 3.6 |
| **Medical insurance** | | | | | | |
| **Private** | 98 | 45.4 | 113 | 50.2 | 211 | 47.8 |
| **Public** | 41 | 19.0 | 43 | 19.1 | 84 | 19.0 |
| **None** | 79 | 36.6 | 79 | 35.1 | 158 | 35.8 |
| **Marital status** | | | | | | |
| **Single** | 62 | 28.7 | 64 | 28.4 | 126 | 28.6 |
| **Married/cohabiting** | 82 | 38.0 | 89 | 39.6 | 171 | 38.8 |
| **Divorce/separated** | 63 | 29.2 | 69 | 30.7 | 132 | 29.9 |
| **Widowed** | 9 | 4.2 | 3 | 1.3 | 12 | 2.7 |
| **Clinical Features** | **n** | **%** | **n** | **%** | **n** | **%** |
| **First episode occurrence before age 18** | 79 | 36.7 | 91 | 40.8 | 170 | 38.8 |
| **Recurrent MDD** | 140 | 68.3 | 156 | 72.2 | 296 | 70.3 |
| **Family history of depression** | 116 | 55.2 | 113 | 50.9 | 229 | 53.0 |
| **Prior suicide attempt** | 45 | 20.8 | 43 | 19.1 | 88 | 20.0 |
| **Duration of current episode $^{3}$ 2 years** | 56 | 26.0 | 62 | 27.8 | 118 | 26.9 |
| | **Mean** | **SD** | **Mean** | **SD** | **Mean** | **SD** |

| | | | | | | |
|---|---|---|---|---|---|---|
| **Age at first episode (years)** | **26.2** | **14.6** | **24.2** | **13.7** | **25.2** | **14.2** |
| **Illness duration (years)** | **14.8** | **12.3** | **18.4** | **13.8** | **16.6** | **13.2** |
| **Number of MDD episodes** | **4.3** | **6.0** | **6.7** | **11.0** | **5.6** | **9.0** |
| **Duration of current episode (months)** | **24.1** | **45.3** | **28.9** | **60.5** | **26.5** | **53.5** |
| **Median duration of current episode (months)** | **8.7** | | **7.8** | | **8.2** | |
| **$HRSD_{17}$ score (at entry into step-2)** | **17.1** | **6.1** | **18.0** | **6.5** | **17.6** | **6.3** |
| **Cumulative Illness Rating Scale** | | | | | | |
| **Categories endorsed** | **2.3** | **1.5** | **2.6** | **1.6** | **2.5** | **1.5** |
| **Total score** | **4.5** | **4.1** | **5.2** | **4.1** | **4.9** | **4.1** |
| **Severity score** | **1.8** | **0.8** | **1.8** | **0.7** | **1.8** | **0.8** |

*** Note that sums do not always equal n due to missing values. Percentages are based on available data.**
